# Time trends, factors associated with, and reasons for COVID-19 vaccine hesitancy in a massive online survey of US adults: January-May 2021

**DOI:** 10.1101/2021.07.20.21260795

**Authors:** Wendy C King, Max Rubinstein, Alex Reinhart, Robin Mejia

**Author notes:** Corresponding author: (RM).

## Abstract

**Importance:** COVID-19 vaccine hesitancy has become a leading barrier to increasing the US vaccination rate.

**Objective:** To evaluate time trends in COVID-19 vaccine intent during the US vaccine rollout, and identify key factors related to and self-reported reasons for COVID-19 vaccine hesitancy in May 2021.

**Design, participants and setting:** A COVID-19 survey was offered to US adult Facebook users in several languages yielding 5,088,772 qualifying responses from January 6 to May 31, 2021. Data was aggregated by month. Survey weights matched the sample to the age, gender, and state profile of the US population.

**Exposure:** Demographics, geographic factors, political/COVID-19 environment, health status, beliefs, and behaviors.

**Main outcome measures:** “If a vaccine to prevent COVID-19 were offered to you today, would you choose to get vaccinated.” Hesitant was defined as responding probably or definitely would not choose to get vaccinated (versus probably or definitely would, or already vaccinated).

**Results:** COVID-19 vaccine hesitancy decreased by one-third from 25.4% (95%CI, 25.3, 25.5) in January to 16.6% (95% CI, 16.4, 16.7) in May, with relatively large decreases among participants with Black, Pacific Islander or Hispanic race/ethnicity and ≤high school education. Independent risk factors for vaccine hesitancy in May (N=525,644) included younger age, non-Asian race, < 4 year college degree, living in a more rural county, living in a county with higher Trump vote share in the 2020 election, lack of worry about COVID-19, working outside the home, never intentionally avoiding contact with others, and no past-year flu vaccine. Differences in hesitancy by race/ethnicity varied by age (e.g., Black adults more hesitant than White adults <35 years old, but less hesitant among adults ≥45 years old). Differences in hesitancy by age varied by race/ethnicity. Almost half of vaccine hesitant respondents reported fear of side effects (49.2% [95%CI, 48.7, 49.7]) and not trusting the COVID-19 vaccine (48.4% [95%CI, 48.0, 48.9]); over one-third reported not trusting the government, not needing the vaccine, and waiting to see if safe. Reasons differed by degree of vaccine intent and by race/ethnicity.

**Conclusion:** COVID-19 vaccine hesitancy varied by demographics, geography, beliefs, and behaviors, indicating a need for a range of messaging and policy options to target high-hesitancy groups.

## Introduction

On December 11, 2020(1), the Federal Drug Administration (FDA) granted the first Emergency Use Authorization of a COVID-19 vaccine in the United States (US) (2). By March 2021, 3 COVID-19 vaccines had been authorized(3), and the president announced procurement of enough doses for every adult to be vaccinated by the end of May 2021 (4). By May 2021, vaccine eligibility was expanded to everyone covered under the FDA authorizations (initially ≥16 years old, expanded to ≥12 years old on May 10 (5)), and efforts to increase vaccine access to underserved populations were underway (6,7). However, COVID-19 vaccine hesitancy (i.e., a refusal or reluctance to be vaccinated) slowed vaccination uptake (8). By July 2021, COVID-19 vaccine hesitancy, which appears to be a distinct phenomenon from general vaccine hesitancy (9), was widely perceived by the public as prolonging the pandemic; on August 1, it was the subject of a New York Times front-page story (10), and was center-stage to disease control discussions as a fourth COVID-19 surge filled hospitals across parts of the US (11,12).

A longitudinal study of US adults (N=7,420) by Daly et al. reported an overall decrease in COVID-19 vaccine hesitancy from 46.0% in October 2020 to 35.2% in March 2021. Throughout this timeframe, younger versus older adults, and Black versus White adults, had greater COVID-19 vaccine hesitancy, however, trends suggested an increase in the age disparity and decrease in the racial disparity in vaccine uptake (13). Understanding how COVID-19 vaccine hesitancy prevalence continued to change up through the time of adult universal eligibility, overall and among subgroups, as well as reasons for hesitancy at the time of universal eligibility is essential for policy makers working to increase vaccination uptake.

Since April 2020, the Delphi Group at Carnegie Mellon University (CMU) has been conducting a national online COVID-19 Trends and Impact Survey (CTIS) (14) in collaboration with a consortium of universities and Facebook Data for Good (14). Among a massive sample of US adults who completed the CTIS January-May, 2021, we report COVID-19 vaccine uptake and intent by month, and evaluate time trends in vaccine hesitancy prevalence by race/ethnicity, education, US region and political environment. For May, the prevalence of COVID-19 vaccine hesitancy is reported by demographics, geographic factors, political/COVID-19 environment, health status, beliefs and behaviors, and associations between each potential risk factor with hesitancy is estimated with and without adjustment for potential confounders. Lastly, we identify the most common reasons for COVID-19 vaccine hesitancy by level of COVID-19 vaccine intent and race/ethnicity.

## Methods

### Survey sampling and weighting

Each month, January-May, 2021, the CTIS was offered to a random sample, stratified by geographic region, of ≈100 million US residents from the Facebook Active User Base who used one of the supported languages (English [American and British], Spanish [Spain and Latin American], French, Brazilian Portuguese, Vietnamese, and simplified Chinese). The offer to participate was shown with a survey link at the top of users’ Facebook News Feed, from once a month to once every six months, depending on their geographic strata, with the intent to yield ≈1.1 million responders monthly, to allow for evaluation of local trends. When individuals clicked through the link, an anonymized unique identifier was generated. CMU returned the unique IDs to Facebook, which created weights that account for the sampling design and non-response; these weights were then post-stratified to match the US general population by age, gender, and state (14,15). The design ensured CMU researchers did not see user names or profile information, and Facebook did not see survey microdata. The CMU Institutional Review Board approved the survey protocol and instrument (STUDY2020_00000162).

### Study sample

The same version of the vaccine uptake and intent questions were offered to all potential CTIS respondents from January 6 to May, 19, 2021, and to approximately 15% of potential respondents from May 20-31. This study is limited to responses from these offers (5,485,862 of 476,648,117; response rate 1.2%). Respondents who did not complete the questions on vaccine uptake and intent (N=365,426), or reported gender as, “prefer to self-describe,” (N=31,664), were excluded, resulting in a sample of 5,088,772; self-described gender (selected by <1% of responders) had a high prevalence of discriminatory descriptions and uncommon responses (e.g., Hispanic ethnicity [41.4%], the oldest age group [23.2% ≥75 years] and highest education level [28.1% Doctorate]), suggesting the survey was not completed in good faith. A sensitivity analysis was conducted including all gender responses.

### Measures

The survey questions and response sets utilized in this report are provided in supporting information, **sAppendix**. Vaccination questions were adapted from CDC-sponsored questions developed for two household panel surveys (16) and shared with us prior to launch. The answer set for reasons for vaccine hesitancy was also expanded through a review of media reports and brainstorming sessions among survey methodologists. We categorized participants as vaccine hesitant if they answered that they probably or definitely would not choose to get vaccinated, “if a vaccine to prevent COVID-19 were offered to you today,” (versus probably or definitely would choose to get vaccinated or were already vaccinated). Those who had already received a COVID-19 vaccine were coded as not hesitant to ensure a consistent study population, as access to vaccinations varied by state and month in the studied timeframe.

The question on gender was developed for this survey; other demographic questions (age, race, ethnicity, education, employment status) were adapted from existing surveys: race and ethnicity from the 2020 Census (17), age categories match the 10-year blocks reported by the American Community Survey (ACS) (18), and education categories were adapted from ACS (19). Participants who reported Hispanic ethnicity were categorized by ethnicity. Non-Hispanic participants were categorized by their race; non-Hispanic participants who reported more than one race were categorized as multi-racial. Participants who did not select one of the named races were categorized as missing. The “less than high school” and “high school graduate or equivalent” categories were combined as “≤high school.” Likewise, “some college” and “2 year degree” were combined as “some college.”

County-level variables were created using participants’ reported ZIP codes: US region (i.e., Census Bureau statistical region (20), dividing West into Mountain and Pacific, as vaccination rates differed in these subregions (21)); urban-rural level of metropolitan statistical area classification (22); and several political/COVID-19 environment indicators: quartile of county COVID-19 death rate in the previous month (April 2021) (23), quartile of county Trump vote share in the 2020 presidential election (calculated as percent voted for Trump minus percent voted for Biden out of total votes within a county; not available for Alaska) (24), and state governor’s political party (not available for US territories) (25).

Questions related to health status, beliefs and behaviors included ever having tested positive for COVID-19, ever diagnosed with specific health conditions (asthma, autoimmune disorder, cancer, chronic obstructive pulmonary disease, diabetes type 1, diabetes type 2, heart disease, high blood pressure, kidney disease, and weakened/compromised immune system), the extent of worry about self or family becoming seriously ill from COVID-19, living with someone age 65 years or older, past-year flu vaccination, and the extent of intentionally avoiding contact with others. Participants were categorized as having no health conditions or at least one health condition. Additionally, participants were categorized as having high blood pressure only, each of the other health conditions with or without high blood pressure, or “multiple conditions,” defined as at least two conditions excluding high blood pressure, which was relatively common and has limited support as a risk-factor for COVID-19 (26).

### Statistical analysis

Data was aggregated by month to evaluate time trends in COVID-19 vaccine uptake and intent. There may have been repeat respondents across months; however, respondents could not be linked longitudinally, so data was treated as repeat cross-sectional surveys. All estimates were generated using survey weights (15). We calculated percentages of COVID-19 vaccine intent or hesitancy by month, as well as first-last month differences, for the full sample, and by race/ethnicity, education, US region, and county Trump support. We limited the race/ethnicity comparison to adults 18-34 years due to an interaction between race/ethnicity and age (reported below with May data).

May 2021 data was used to assess how demographics, geographic factors, political/COVID-19 environment, health status, beliefs and behaviors related to COVID-19 vaccine hesitancy. Specifically, we calculated percentages of COVID-19 vaccine hesitancy by all covariates and used a series of weighted Poisson regression models with robust error variance to estimate the risk ratios (RR) for vaccine hesitancy for each variable (27,28). Adjusted risk ratios (aRR) were estimated from a single Poisson regression model including all covariates and an interaction term for age group and race/ethnicity. To enable model fitting with an interaction term for age and race, where age data was missing but race/ethnicity was available, race/ethnicity was recoded to missing in the May sample. This affected 70 Hispanics, 262 Whites, 46 Blacks, 13 Asians, 15 Native Americans, 1 Pacific Islanders, and 30 multi-racial respondents (0.08% of our sample). In a second multivariable model, a simplified health conditions variable (none, at least one, described above) was replaced with the version specifying specific conditions to estimate aRR by health condition. We calculated percentages of participants with each reason for hesitancy by 3 levels of vaccine intent (definitely no, probably no, and probably yes), and by race/ethnicity among hesitant participants.

For explanatory variables, “missing” was treated as a response category in the analyses. For all parameters, 95% confidence intervals (CI) were calculated using robust standard errors (29). Analyses were conducted in R (Version 4.0.2, R Core Team, Vienna, Austria).

## Results

Study flow overall and by month is provided in supporting information, **sTable 1**. The January, February, March, April and May samples had 1,195,650; 1,142,195; 1,209,536; 1,015,747; and 525,644 participants, respectively.

### Participant characteristics

Excluding missing responses, weighted May participants had a median age range of 55-64 years; 45.7% identified as male, 53.2% female, 1.1% nonbinary; 16.4% were Hispanic, 69.1 % White, 6.5% Black, 3.6% Asian, 0.8% Native American, 0.24% Pacific Islander, and 3.3% were multi-racial; 22.5% had ≤high school education; 41.0% ≥four-year college degree. Over half (56.0%) worked for pay; 42.9% worked outside the home. Demographics were similar January through April (**sTable2**).

### COVID-19 vaccine receipt and intent over time

Hesitancy decreased each month, with a one-third decrease from 25.4% (95%CI 25.3,25.5) in January to 16.6% (95% CI, 16.4,16.7) in May, 2021. There was a bigger decrease in percentage points in the response “probably not” (−7.1%[95% CI -7.2, -6.9]) versus “definitely not” (−1.8% [95% CI -1.9, -1.7]) (**Figure 1; sTable 3**).

**Figure 1.**
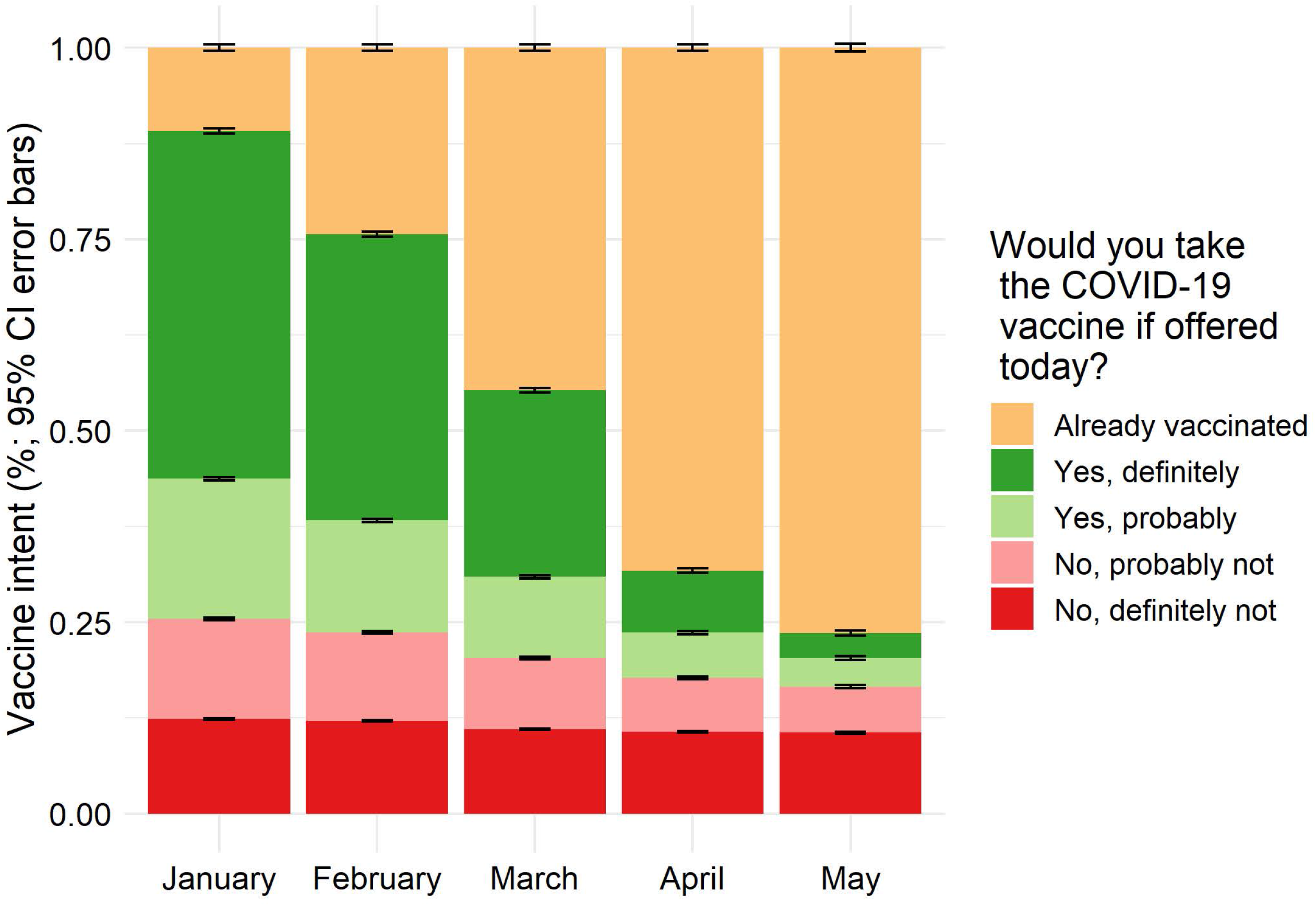
COVID-19 vaccine receipt and intent among US adults by month (January-May, 2021) Vaccine hesitancy decreased among adults each month from January to May, mostly due to a decrease in the response, “probably not” (−7.1 percentage points (%) [95% CI -7.2, -6.9]) versus “definitely not” (−1.8 % [95% CI -1.9, -1.7]).

Per **Figure 2**, from January to May the gap in percent hesitant between race/ethnicity groups among younger adults (**panel A**) and education levels among all respondents (**panel B**) decreased, with the biggest decreases among two of the three most hesitant race/ethnicity groups (e.g., Black and Pacific Islander but not Native American) and the two most hesitant education groups (≤high school and some college education) in January. Hesitancy appeared relatively constant among those with a professional degree (e.g. MD, JD) or Doctorate. Decreases in percent hesitant over time were fairly similar across US regions (**panel C**), with a slightly smaller decrease in the Mountain region and slightly larger decrease in the South. The gap in percent hesitant by Trump vote share increased slightly from January to May, with the highest quartile, which was the most hesitant group, having the smallest decrease (**panel D**). Supporting data is provided in **sTable 4**.

**Figure 2.**
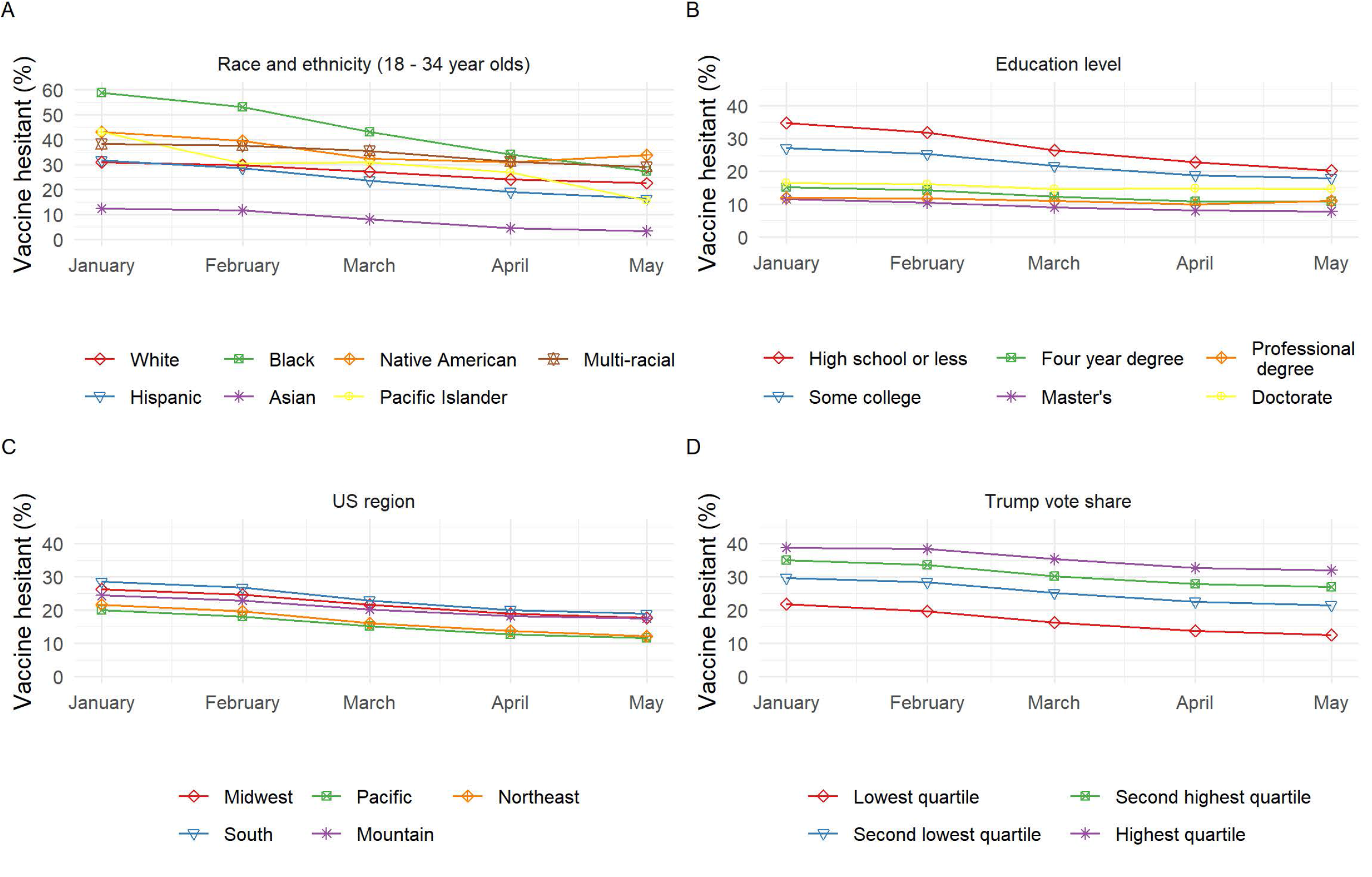
COVID-19 vaccine hesitancy by race/ethnicity (ages 18-34 years^a^), education level, US region and county Trump vote share in the 2020 presidential election among US adults by month (January-May, 2021) Between January and May, the gap in percent hesitant between race/ethnicity groups among adults 18-34 years (panel A) and education levels among all ages (panel B) decreased, with the biggest decreases among the most hesitant groups (e.g., Black race and ≤high school education, respectively). Changes in percent hesitant over time were fairly similar across US regions (panel C); however, there was a slightly smaller decrease in the Mountain region and slightly larger decrease in the South versus other regions. The gap in percent hesitant by county political environment, quantified in quartiles of percent Trump vote share in the 2020 presidential election, increased slightly between January and May, with the most hesitant group (highest quartile) having the smallest decrease (panel D). ^a^ There was a significant interaction between race/ethnicity group and age group. Vaccine hesitancy for all race/ethnicity groups by all age groups is provided in **sTable 6**.

### Factors related to COVID-19 vaccine hesitancy

Hesitancy in May, 2021 is reported by participant demographics and geographic factors in **Table 1** (N=525,644). Although hesitancy was lower in females versus males (RR 0.79, 95% CI 0.78, 0.81), with adjustment for covariates (i.e., variables reported in **Tables 1** and **2**), female gender was associated with higher hesitancy (aRR 1.12, 95%CI 1.10, 1.14). Non-binary adults had similar hesitancy to males (RR 1.10, 95%CI, 0.97, 1.22; aRR 0.99, 95%CI 0.88, 1.10).

**Table 1.** COVID-19 vaccine hesitancy in May 2021 by demographics among US adults (N=525,644)

|  | Sample |  | COVID-19 vaccine hesitant |  |  |
| --- | --- | --- | --- | --- | --- |
|  | n | % | % (95% CI) | RR (95% CI)<sup>b</sup> | Adj. RR (95% CI)<sup>c</sup> |
| **Gender** |  |  |  |  |  |
| Male | 159427 | 30.3 | 16.6 (16.4, 16.9) | 1.0 (NA) | 1.0 (NA) |
| Female | 294983 | 56.1 | 13.2 (13.1, 13.4) | 0.79 (0.78, 0.81) | 1.12 (1.10, 1.14) |
| Non-binary | 3232 | 0.6 | 18.2 (16.1, 20.3) | 1.10 (0.97, 1.22) | 0.99 (0.88, 1.10) |
| No response | 68002 | 12.9 | 26.3 (25.8, 26.7) | 1.58 (1.54, 1.61) | 1.39 (1.34, 1.44) |
| **Age group** |  |  |  |  |  |
| 18-24 years | 15382 | 2.9 | 22.1 (21.3, 23.0) | 2.76 (2.64, 2.88) | <sup>d</sup> |
| 25-34 years | 52015 | 9.9 | 20.5 (20.1, 20.9) | 2.56 (2.48, 2.64) |  |
| 35-44 years | 72541 | 13.8 | 17.8 (17.5, 18.2) | 2.22 (2.16, 2.29) |  |
| 45-54 years | 81005 | 15.4 | 16.6 (16.3, 16.8) | 2.07 (2.01, 2.13) |  |
| 55-64 years | 102934 | 19.6 | 12.7 (12.4, 12.9) | 1.58 (1.53, 1.63) |  |
| 65-74 years | 95607 | 18.2 | 8.0 (7.8, 8.2) | 1.0 (NA) |  |
| ≥ 75 years | 41799 | 8.0 | 6.9 (6.6, 7.2) | 0.86 (0.82, 0.90) |  |
| Missing | 64361 | 12.2 | 24.4 (23.9, 24.8) | 3.04 (2.95, 3.13) |  |
| **Race/ethnicity<sup>a</sup>** |  |  |  |  |  |
| White | 337260 | 64.2 | 15.5 (15.4, 15.7) | 1.0 (NA) | <sup>d</sup> |
| Hispanic | 56333 | 10.7 | 11.9 (11.5, 12.3) | 0.77 (0.74, 0.79) |  |
| Black | 28546 | 5.4 | 12.9 (12.4, 13.4) | 0.83 (0.79, 0.86) |  |
| Asian | 11962 | 2.3 | 2.9 (2.5, 3.3) | 0.19 (0.16, 0.21) |  |
| Native American | 3944 | 0.8 | 24.9 (23.0, 26.8) | 1.60 (1.48, 1.73) |  |
| Pacific Islander | 992 | 0.2 | 13.5 (10.9, 16.0) | 0.87 (0.70, 1.03) |  |
| Multi-racial | 13009 | 2.5 | 26.9 (25.9, 27.9) | 1.73 (1.67, 1.80) |  |
| Missing | 73598 | 14.0 | 25.9 (25.4, 26.3) | 1.67 (1.63, 1.70) |  |
| **Education level** |  |  |  |  |  |
| ≤ High school | 91859 | 17.5 | 20.3 (20.0, 20.7) | 1.89 (1.84, 1.94) | 1.58 (1.54, 1.62) |
| Some college | 166070 | 31.6 | 17.9 (17.7, 18.2) | 1.67 (1.62, 1.71) | 1.38 (1.35, 1.41) |
| 4 year college | 110440 | 21.0 | 10.8 (10.5, 11.0) | 1.0 (NA) | 1.0 (NA) |
| Master's | 62503 | 11.9 | 7.8 (7.6, 8.1) | 0.73 (0.70, 0.76) | 0.88 (0.85, 0.91) |
| Professional (e.g., MD, JD) | 14733 | 2.8 | 11.1 (10.4, 11.8) | 1.03 (0.96, 1.10) | 1.08 (1.02, 1.14) |
| Doctorate | 9975 | 1.9 | 14.6 (13.5, 15.6) | 1.35 (1.25, 1.45) | 1.20 (1.14, 1.26) |
| Missing | 70064 | 13.3 | 23.7 (23.3, 24.1) | 2.20 (2.14, 2.27) | 1.19 (1.10, 1.27) |
| **Employment status** |  |  |  |  |  |
| Work outside home | 174011 | 33.1 | 20.2 (19.9, 20.4) | 2.44 (2.35, 2.53) | 1.33 (1.29, 1.38) |
| Work at home | 57015 | 10.8 | 8.3 (8.0, 8.6) | 1.0 (NA) | 1.0 (NA) |
| Does not work for pay | 221728 | 42.2 | 12.4 (12.2, 12.6) | 1.50 (1.44, 1.55) | 1.35 (1.30, 1.40) |
| Missing | 72890 | 13.9 | 23.6 (23.2, 24.1) | 2.86 (2.75, 2.97) | 1.35 (1.26, 1.43) |
| **US Region** |  |  |  |  |  |
| Midwest | 125878 | 23.9 | 17.7 (17.4, 17.9) | 1.51 (1.47, 1.56) | 1.10 (1.07, 1.13) |
| South | 181704 | 34.6 | 18.8 (18.6, 19.1) | 1.62 (1.56, 1.67) | 1.14 (1.11, 1.17) |
| Pacific | 72997 | 13.9 | 11.7 (11.3, 12.0) | 1.0 (NA) | 1.0 (NA) |
| Mountain | 41975 | 8.0 | 17.5 (17.0, 18.0) | 1.50 (1.44, 1.57) | 1.12 (1.08, 1.16) |
| Northeast | 87740 | 16.7 | 12.2 (11.9, 12.5) | 1.05 (1.01, 1.09) | 0.96 (0.93, 0.99) |
| Territories | 190 | 0.0 | 10.7 (5.5, 15.9) | 0.92 (0.47, 1.37) | 0.64 (0.41, 0.86) |
| Missing | 15160 | 2.9 | 29.8 (28.8, 30.7) | 2.55 (2.44, 2.66) | <sup>e</sup> |
| **County urban classification** |  |  |  |  |  |
| Large central metro | 119939 | 22.8 | 11.4 (11.2, 11.7) | 1.0 (NA) | 1.0 (NA) |
| Large fringe metro | 115240 | 21.9 | 13.9 (13.6, 14.1) | 1.22 (1.18, 1.25) | 1.03 (1.00, 1.06) |
| Medium metro | 137628 | 26.2 | 16.4 (16.1, 16.7) | 1.44 (1.40, 1.48) | 1.13 (1.10, 1.16) |
| Small metro | 57380 | 10.9 | 20.5 (20.0, 20.9) | 1.80 (1.74, 1.85) | 1.18 (1.14, 1.22) |
| Micropolitan | 48869 | 9.3 | 23.6 (23.1, 24.1) | 2.07 (2.00, 2.13) | 1.20 (1.16, 1.23) |
| Non-core | 31238 | 5.9 | 26.9 (26.3, 27.6) | 2.36 (2.28, 2.44) | 1.23 (1.19, 1.28) |
| Missing | 15350 | 2.9 | 29.5 (28.6, 30.5) | 2.59 (2.49, 2.69) | <sup>e</sup> |

Juris Doctorate= JD; MD=Doctor of Medicine; NA=not applicable; NH=Non-Hispanic
<sup>a</sup> Race/ethnicity groups other than the group labeled “Hispanic” are non-Hispanic.
<sup>b</sup> A series of weighted Poisson regression models with robust error variance were used to estimate the risk ratios.
<sup>c</sup> Adjusted risk ratios were estimated from a single Poisson regression model including all covariates and an interaction term for age group and race/ethnicity.
<sup>d</sup> Due to an interaction between age group and race/ethnicity, adjusted relative risks from the multivariable model are reported in supporting information (age group stratified by race in sTable 5; race/ethnicity by age group in sTable 6).
<sup>e</sup> Reliable estimates could not be calculated for the missing category for variables based on participants’ zip code, due to collinearity.

In general, younger age and non-Asian race (particularly multi-racial and Native American), were related to greater hesitancy. However, an interaction was observed between age and race/ethnicity categories (**Figure 3**). Differences in hesitancy by age were more pronounced in Blacks and less pronounced in Asians and Whites. Differences in hesitancy by race/ethnicity were more pronounced in younger adults and adults ≥75 years. Furthermore, for some comparisons, the direction of the difference differed by age. For example, for Blacks versus Whites, the RR of hesitancy was 1.31 (95%CI, 1.12, 1.51) in 18-24 year olds, versus 0.50 (95%CI, 0.34, 0.67) in ≥75 year olds. **sTables 5** and **6** provide RRs for age groups stratified by race/ethnicity groups and vice versa, with and without adjustment for all covariates. Age and race/ethnicity differences were generally attenuated in the full multivariable model, but still present.

**Figure 3.**
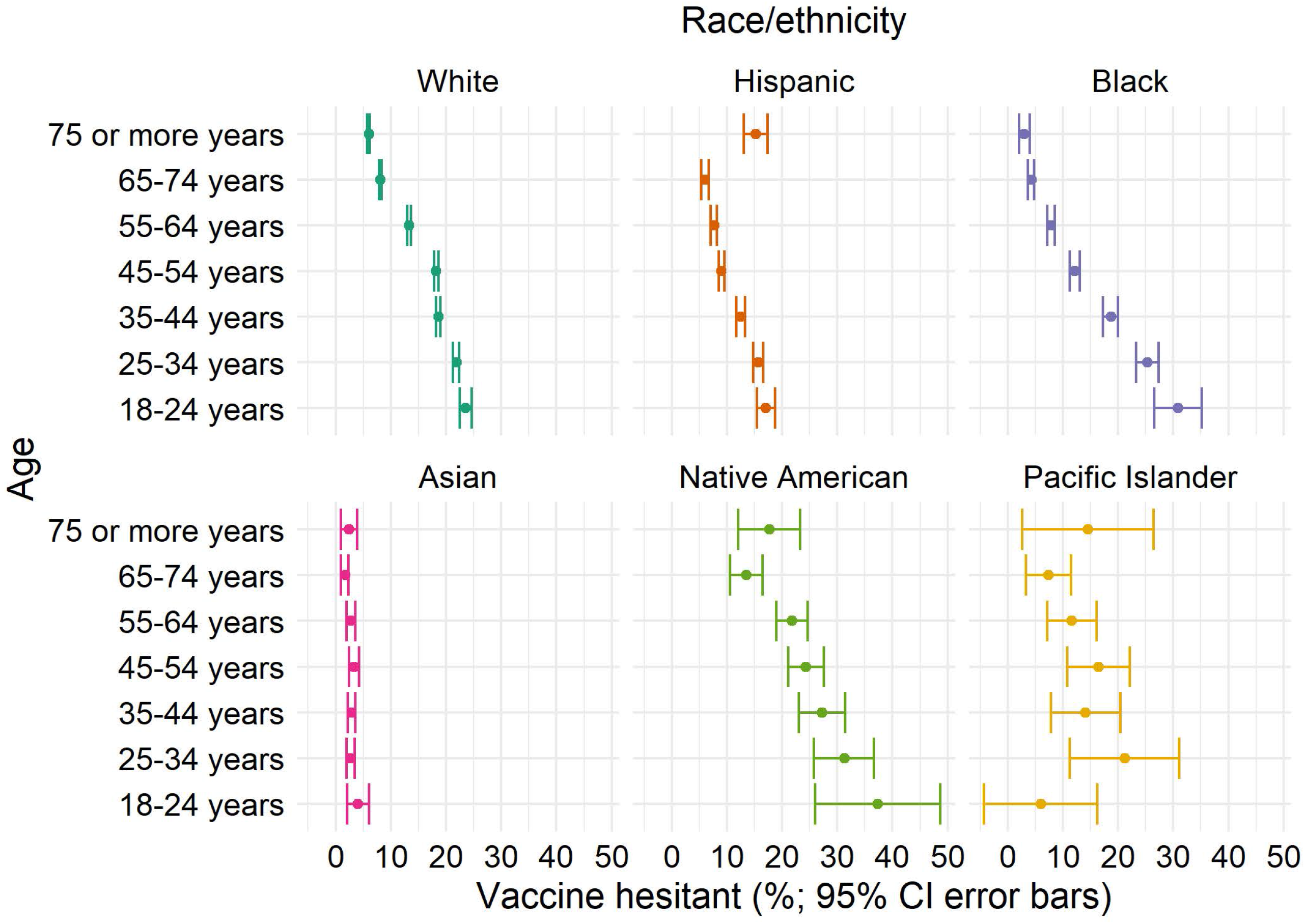
COVID-19 vaccine hesitancy by age group, stratified by race/ethnicity, among US adults, May 2021 Differences in hesitancy by age (e.g., 18-24 year-olds versus 65-74 year-olds) were more pronounced in Blacks (RR=7.26 [95%CI, 5.92, 8.61]) versus Whites (RR=2.89 [95% CI 2.75, 3.04]) or other race/ethnicity groups. Asians had hesitancy <5% hesitancy in all age groups. Differences in hesitancy by race/ethnicity were more pronounced in younger adults and adults ≥75 years.

The association between hesitancy and education level followed a U-shaped curve with the lowest hesitancy among those with a master’s degree, followed by those with a 4 year college degree, then a professional degree, and a doctorate. The highest hesitancy was among those with ≤high school education or some college (RR=1.89 [95%CI 1.84, 1.94] and 1.67 [95%CI 1.62, 1.71], respectively, versus a 4 year college degree). Additional demographic risk factors for hesitancy included working outside the home (RR=2.44 [95%CI 2.35, 2.53]) or not working for pay (RR=1.50 [95% CI: 1.44, 1.55]) versus working at home; living in the South (RR=1.62 95%CI 1.56, 1.67]), Midwest (RR=1.51 [95%CI 1.47, 1.56]) or Mountain (RR=1.50 [95%CI 1.44, 1.57]) versus the Pacific US region; and in a less urban county (e.g., RR=2.36 [95 CI, 2.28, 2.44] for non-core versus large central metro). Associations were attenuated with adjustment (**Table 1**).

COVID-19 vaccine hesitancy is reported by indicators of political/COVID-19 environment, health status, and beliefs and behaviors in **Table 2**. Risk factors for hesitancy were living in a state with Republican versus Democratic governor (RR=1.34 [95%CI 1.32, 1.36]), living in a county with higher Trump support (e.g., RR=2.57 [95%CI 2.50, 2.65] for highest versus lowest quartile), living in a county with a lower April COVID-19 death rate (e.g., RR=0.70 [95%CI 0.68, 0.73] for highest versus lowest quartile), history of a positive COVID-19 test versus no history (RR=1.26 [95%CI 1.23, 1.29]), not having versus having a high-risk health condition (RR=1.41 [95%CI 1.38, 1.43]), less worry about self or immediate family becoming seriously ill from COVID-19 (e.g., RR=3.66 [3.58, 3.74] for not worried at all versus worried), not having versus having received a past-year flu vaccine (RR=4.97 [95%CI 4.85, 5.08]), and not avoiding versus avoiding contact with others (e.g., RR=3.96 [95%CI 3.84, 4.07] for none versus all of the time). Political affiliation of state governor was excluded from the multivariable model due to collinearity with county Trump support. Associations from the multivariable model were attenuated but in the same direction, except for April 2021 COVID-19 death rate, which was not independently related to hesitancy.

**Table 2.** COVID-19 vaccine hesitancy in May 2021 by political/ COVID-19 environment, health status, beliefs and behaviors among US adults (N=525,644)

|  | Sample |  | COVID-19 vaccine hesitant |  |  |
| --- | --- | --- | --- | --- | --- |
|  | N | % | % (95% CI) | RR (95% CI)<sup>a</sup> | Adj. RR (95% CI)<sup>b</sup> |
| **State governor's political party** |  |  |  |  |  |
| Democratic | 280671 | 53.4 | 14.0 (13.8, 14.2) | 1.0 (NA) | <sup>c</sup> |
| Republican | 228790 | 43.5 | 18.8 (18.6, 19.0) | 1.34 (1.32, 1.36) |  |
| Missing | 16183 | 3.1 | 28.4 (27.4, 29.3) | 2.03 (1.95, 2.10) |  |
| **County Trump vote total minus Biden vote total in 2020 presidential election** |  |  |  |  |  |
| Lowest quartile | 341190 | 64.9 | 12.4 (12.3, 12.6) | 1.0 (NA) | 1.0 (NA) |
| Second lowest quartile | 100974 | 19.2 | 21.4 (21.1, 21.8) | 1.73 (1.69, 1.76) | 1.28 (1.25, 1.30) |
| Second highest quartile | 47061 | 9.0 | 27.0 (26.5, 27.6) | 2.18 (2.12, 2.23) | 1.35 (1.31, 1.38) |
| Highest quartile | 19531 | 3.7 | 31.9 (31.1, 32.8) | 2.57 (2.50, 2.65) | 1.44 (1.40, 1.49) |
| Missing | 16888 | 3.2 | 29.0 (28.1, 29.9) | 2.34 (2.25, 2.42) | 1.15 (1.02, 1.28) |
| **County COVID-19 April 2021 county death rate** |  |  |  |  |  |
| Lowest quartile | 25972 | 4.9 | 23.8 (23.2, 24.5) | 1.0 (NA) | - |
| Second lowest quartile | 167874 | 31.9 | 15.8 (15.5, 16.0) | 0.66 (0.64, 0.68) | 0.97 (0.94, 1.00) |
| Second highest quartile | 213276 | 40.6 | 15.3 (15.1, 15.5) | 0.64 (0.62, 0.66) | 1.00 (0.97, 1.03) |
| Highest quartile | 103140 | 19.6 | 16.7 (16.4, 17.0) | 0.70 (0.68, 0.73) | 1.01 (0.98, 1.04) |
| Missing | 15382 | 2.9 | 29.5 (28.5, 30.5) | 1.24 (1.18, 1.29) | <sup>d</sup> |
| **Ever tested positive for COVID-19** |  |  |  |  |  |
| Yes | 55306 | 10.5 | 20.2 (19.8, 20.6) | 1.26 (1.23, 1.29) | 1.11 (1.09, 1.14) |
| No or unsure | 467169 | 88.9 | 16.1 (15.9, 16.2) | 1.0 (NA) | 1.0 (NA) |
| No response | 3169 | 0.6 | 17.5 (15.7, 19.2) | 1.09 (0.98, 1.20) | 0.91 (0.83, 0.99) |
| **Ever diagnosed with high-risk medical condition** |  |  |  |  |  |
| One or more conditions | 322188 | 61.3 | 13.3 (13.2, 13.5) | 1.0 (NA) | 1.0 (NA) |
| No conditions | 182782 | 34.8 | 18.7 (18.5, 19.0) | 1.41 (1.38, 1.43) | 1.00 (0.99, 1.02) |
| Missing | 20674 | 3.9 | 35.5 (34.6, 36.3) | 2.66 (2.59, 2.74) | 1.71 (1.66, 1.76) |
| **Extent worried that you or someone in immediate family might become seriously ill from COVID-19** |  |  |  |  |  |
| Worried | 209073 | 39.8 | 8.7 (8.6, 8.9) | 1.0 (NA) | 1.0 (NA) |
| Not too worried | 164156 | 31.2 | 13.5 (13.3, 13.7) | 1.55 (1.51, 1.59) | 1.31 (1.28, 1.34) |
| Not worried at all | 96414 | 18.3 | 31.9 (31.5, 32.3) | 3.66 (3.58, 3.74) | 1.79 (1.75, 1.83) |
| Missing | 56001 | 10.7 | 24.7 (24.2, 25.2) | 2.84 (2.76, 2.92) | 1.31 (1.11, 1.52) |
| **Lives with someone or is 65 years or older, including self** |  |  |  |  |  |
| Yes | 207265 | 39.4 | 11.6 (11.4, 11.8) | 1.0 (NA) |  |
| No | 203619 | 38.7 | 17.7 (17.5, 17.9) | 1.53 (1.49, 1.56) | 1.08 (1.05, 1.11) |
| No response | 114760 | 21.8 | 20.9 (20.6, 21.2) | 1.80 (1.76, 1.84) | 1.11 (1.09, 1.14) |
| **Past-year flu vaccine** |  |  |  |  |  |
| Yes | 279745 | 53.2 | 5.5 (5.4, 5.6) | 1.0 (NA) | 1.0 (NA) |
| No or unsure | 190331 | 36.2 | 27.2 (26.9, 27.5) | 4.97 (4.85, 5.08) | 3.27 (3.19, 3.35) |
| Missing | 55568 | 10.6 | 24.8 (24.3, 25.3) | 4.53 (4.40, 4.66) | 2.11 (1.76, 2.46) |
| **Extent intentionally avoiding contact with others** |  |  |  |  |  |
| All of the time | 66728 | 12.7 | 10.8 (10.5, 11.1) | 1.0 (NA) | - |
| Most of the time | 141838 | 27.0 | 8.4 (8.2, 8.6) | 0.77 (0.75, 0.80) | 0.88 (0.85, 0.91) |
| Some of the time | 186720 | 35.5 | 8.9 (8.7, 9.0) | 0.82 (0.79, 0.85) | 0.89 (0.86, 0.92) |
| None of the time | 83338 | 15.9 | 42.8 (42.4, 43.3) | 3.96 (3.84, 4.07) | 2.45 (2.37, 2.53) |
| Missing | 47020 | 8.9 | 26.1 (25.5, 26.6) | 2.41 (2.33, 2.49) | 1.46 (1.38, 1.55) |

NA=not applicable
<sup>a</sup> A series of weighted Poisson regression models with robust error variance were used to estimate the risk ratios.
<sup>b</sup> Adjusted risk ratios were estimated from a single Poisson regression model including all covariates and an interaction term for age group and race/ethnicity.
<sup>c</sup> State governor's political party was excluded from the multivariable model due to collinearity with county Trump vote share.
<sup>d</sup> Reliable estimates could not be calculated for the missing category for variables based on participants' zip code

COVID-19 vaccine hesitancy by specific health conditions is provided in **sTable 7**. Compared to participants reporting none of the queried high-risk health conditions, hesitancy was lower among participants with each health condition category except weakened or compromised immune system (RR 1.10, [95%CI 1.02, 1.19]; aRR 1.43 [95%CI 1.33, 1.52]).

A sensitivity analysis including participants selecting “prefer to self-describe” gender is provided in **sTable 8** and **sTable 9**. Overall, results were similar; however, hesitancy prevalence was higher for a few categories (e.g., age ≥75 years, Hispanic, and Doctorate) where mis-reporting was suspected.

### Reasons for COVID-19 vaccine hesitancy

Reasons for hesitancy by applicable levels of intent are reported in **Table 3**. Concern about side effects was chosen most frequently at 49.2% (95%CI, 48.7, 49.7) among hesitant participants, and was similarly common across intent levels. In contrast, among adults who would “definitely not” choose to be vaccinated, not trusting the COVID-19 vaccine and not trusting the government were reported most frequently (59.2% [95%CI, 58.6, 59.8] and 51.2% [95%CI, 50.6, 51.7], respectively), double the prevalence seen among those who would “probably not” get vaccinated and almost quadruple that of adults who “yes, probably” would. Conversely, 52.3% (95%CI, 51.5, 53.1) of the “probably not” group said they would wait to see if it was safe, versus only 24.3% (95%CI, 23.8, 24.8) of the “definitely not” group. Compared to most reasons for vaccine hesitancy, not liking vaccines in general was chosen less frequently across all intent levels (7.6% in “probably yes” to 17.6% in “definitely not”).

**Table 3.**
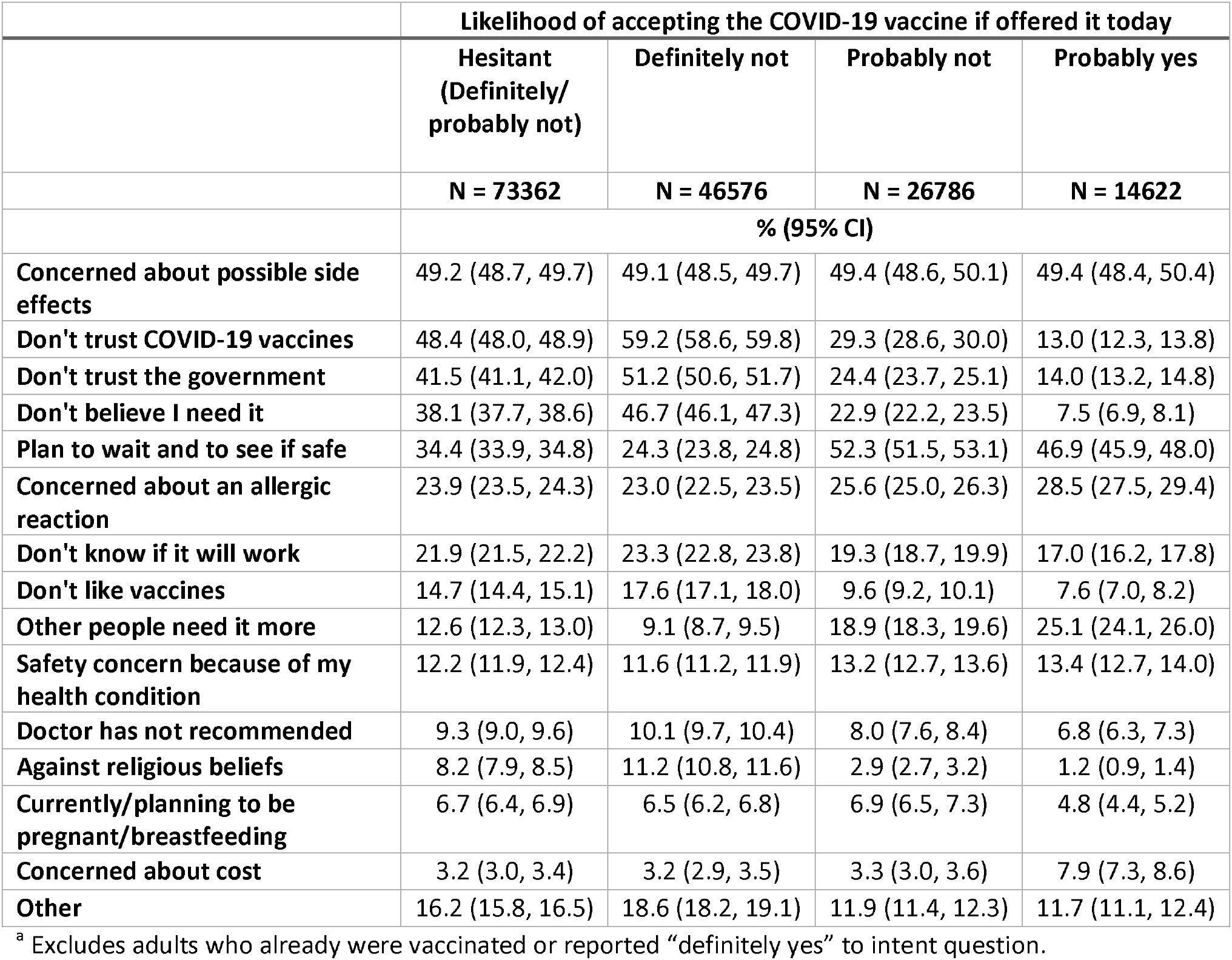
Reasons for not getting the COVID-19 vaccine in May, 2021, by vaccine intent level among US adults (N=87,984)^a^

|  | Likelihood of accepting the COVID-19 vaccine if offered it today |  |  |  |
| --- | --- | --- | --- | --- |
|  | Hesitant<br>(Definitely/<br>probably not) | Definitely not | Probably not | Probably yes |
|  | N = 73362 | N = 46576 | N = 26786 | N = 14622 |
|  | % (95% CI) |  |  |  |
| Concerned about possible side effects | 49.2 (48.7, 49.7) | 49.1 (48.5, 49.7) | 49.4 (48.6, 50.1) | 49.4 (48.4, 50.4) |
| Don't trust COVID-19 vaccines | 48.4 (48.0, 48.9) | 59.2 (58.6, 59.8) | 29.3 (28.6, 30.0) | 13.0 (12.3, 13.8) |
| Don't trust the government | 41.5 (41.1, 42.0) | 51.2 (50.6, 51.7) | 24.4 (23.7, 25.1) | 14.0 (13.2, 14.8) |
| Don't believe I need it | 38.1 (37.7, 38.6) | 46.7 (46.1, 47.3) | 22.9 (22.2, 23.5) | 7.5 (6.9, 8.1) |
| Plan to wait and to see if safe | 34.4 (33.9, 34.8) | 24.3 (23.8, 24.8) | 52.3 (51.5, 53.1) | 46.9 (45.9, 48.0) |
| Concerned about an allergic reaction | 23.9 (23.5, 24.3) | 23.0 (22.5, 23.5) | 25.6 (25.0, 26.3) | 28.5 (27.5, 29.4) |
| Don't know if it will work | 21.9 (21.5, 22.2) | 23.3 (22.8, 23.8) | 19.3 (18.7, 19.9) | 17.0 (16.2, 17.8) |
| Don't like vaccines | 14.7 (14.4, 15.1) | 17.6 (17.1, 18.0) | 9.6 (9.2, 10.1) | 7.6 (7.0, 8.2) |
| Other people need it more | 12.6 (12.3, 13.0) | 9.1 (8.7, 9.5) | 18.9 (18.3, 19.6) | 25.1 (24.1, 26.0) |
| Safety concern because of my health condition | 12.2 (11.9, 12.4) | 11.6 (11.2, 11.9) | 13.2 (12.7, 13.6) | 13.4 (12.7, 14.0) |
| Doctor has not recommended | 9.3 (9.0, 9.6) | 10.1 (9.7, 10.4) | 8.0 (7.6, 8.4) | 6.8 (6.3, 7.3) |
| Against religious beliefs | 8.2 (7.9, 8.5) | 11.2 (10.8, 11.6) | 2.9 (2.7, 3.2) | 1.2 (0.9, 1.4) |
| Currently/planning to be pregnant/breastfeeding | 6.7 (6.4, 6.9) | 6.5 (6.2, 6.8) | 6.9 (6.5, 7.3) | 4.8 (4.4, 5.2) |
| Concerned about cost | 3.2 (3.0, 3.4) | 3.2 (2.9, 3.5) | 3.3 (3.0, 3.6) | 7.9 (7.3, 8.6) |
| Other | 16.2 (15.8, 16.5) | 18.6 (18.2, 19.1) | 11.9 (11.4, 12.3) | 11.7 (11.1, 12.4) |

<sup>a</sup> Excludes adults who already were vaccinated or reported "definitely yes" to intent question.

Reasons for COVID-19 vaccine hesitancy among hesitant US adults by race/ethnicity are provided in **sTable 10**. Concern about side effects, followed by not trusting the COVID-19 vaccine, were the most common concerns in all race/ethnicity groups, with the ranking reversed among Native Americans and Pacific Islanders. Both were chosen by more than 40% of each group except Asians (38.4% [95%CI, 31.5, 45.4] lacked trust in COVID-19 vaccines). There was more racial variability in not trusting the government (range: 26.2-50.0%; >40% among multi-racial, White, and Native American); in waiting to see if safe (range: 27.0-42.5%; >40% among Asian, Hispanic, and Black), and in do not need (range: 22.3-47.0%; >40% among multi-racial and White). Other reasons were chosen by <40% of hesitant adults in each race/ethnicity group.

## Discussion

In this massive national survey of US adults, COVID-19 vaccine hesitancy decreased by one-third between January and May, 2021. However, there was minimal change in the prevalence of the most hesitant subgroup (those responding “definitely not”). Additionally, while a decrease in hesitancy was observed across almost all categories evaluated, there was a substantial difference in the magnitude of change of education and race/ethnicity categories. The largest decreases were seen in the race and education categories with the highest January hesitancy prevalence (e.g., Black and Pacific Islander race, ≤high school and some college education), such that the disparity in hesitancy by these factors decreased. Still, May 2021 data indicates that at the launch of universal vaccine eligibility, COVID-19 vaccine hesitancy varied by a wide array of demographic, geographic, political and COVID-19 environment, health, belief and behavioral factors. Additionally, important differences in reasons for COVID-19 vaccine hesitancy exist both by degree of vaccine intent and racial/ethnic groups.

Racial/ethnic disparities have been observed in all aspects of the COVID-19 pandemic(30), with communities of color experiencing higher rates of SARS-CoV-2 infection, as well as COVID-19-related hospitalizations and mortality (31,32). Racial/ethnic disparities in COVID-19 vaccine acceptance at the start of the vaccine rollout threatened to continue this trend (33), with unclear/unreliable information on COVID-19 vaccines, concerns about research ethics, and access barriers contributing to COVID-19 vaccine hesitancy (34). In response, groups from at-risk communities initiated targeted outreach campaigns (35). The large decreases in COVID-19 vaccine hesitancy we measured among Blacks and Pacific Islanders, suggest that messaging and outreach campaigns, combined with time to observe the initial months of the vaccine rollout, had positive effects; by May 2021, Blacks and Pacific Islanders joined Asian and Hispanics in having a lower prevalence of hesitancy than Whites. However, among younger adults, Blacks, Native Americans, and Multi-racial groups continued to be the most hesitant. Experts continue to recommend targeted campaigns to overcome structural barriers on racial and ethnic disparities in COVID-19 vaccine uptake (36). Our study indicates that in May, 2021, Black and Hispanic adults were more likely to be concerned about safety compared to White adults, while White adults were more likely to report not trusting the government and not needing the vaccine than Black and Hispanic adults.

January through May, there was a dose-response relationship between relative degree of local Trump support in the 2020 presidential election and COVID-19 vaccine hesitancy that grew over time; by May, even after controlling for potential confounders, those living in a county in the top quartile were 44% more at risk of being hesitant, highlighting the politicization of public health recommendations.

Those who were not intentionally avoiding contact with others had much higher likelihood of COVID-19 vaccine hesitancy, as did those working outside the home, indicating non-vaccinated individuals may be the most likely to engage in activities with transmission risk, and thus, are worthy of further study and focused vaccination uptake efforts.

In May 2021, only 14.7% of COVID-19 vaccine hesitant respondents chose not liking vaccines as a reason, indicating that COVID-19 vaccine hesitancy is likely a distinct phenomenon from general vaccine hesitancy among US adults in general, as well as employed adults(9). However, we also found that those who had not received a flu vaccine were 3.3 times more likely to be COVID-19 vaccine hesitant, even after controlling for a wide array of covariates, which indicates that efforts to increase flu vaccination uptake should address reasons of COVID-19 vaccine hesitancy and vice versa.

Concern about side effects of COVID-19 vaccines was common across levels of vaccine intent and among all racial groups. However, several reasons for COVID-19 vaccine hesitancy varied substantially by subgroups. Among less hesitant (“probably not”) participants, waiting to see if the vaccine is safe was a common response, suggesting messaging about safety and policy interventions to address downstream impacts of vaccine side effects, such as potential lost work, could be impactful. Conversely, the most hesitant (“definitely not”) participants commonly reported they don’t trust the COVID-19 vaccine, don’t trust the government, and/or don’t need the vaccine, suggesting the existence of a subgroup with entrenched hesitancy and high distrust that may only respond to vaccine mandates.

### Limitations and strengths

The study employs a novel sampling method with a soft ask and low response rate, the effect of which has not yet been fully studied. Survey weights adjust for non-response and coverage bias (i.e., matching the sample to gender, age, and geographic profile of the US). However, Facebook users may differ from non-users, and our sample is more educated (37) and has higher vaccine uptake(8) than the general population, indicating that we underestimated vaccine hesitancy compared to the general US population. Importantly, CTIS results have been consistent over time, follow similar patterns observed by others (13), and have been used to track trends and inform policies (38,39).

Demographic questions were asked at the end of the survey and had high unit non-response (e.g., 12% for age). To maximize the analysis sample and control for bias, “missing” was included as a variable category; however, interpretation of estimates for this category is difficult. Additionally, we assume the survey was completed in good faith. However, as noted above, a small percentage of participants selected “prefer to self-describe” gender to make discriminatory statements and the frequency of other characteristics in this group was suspect. Thus, they were excluded from the primary analysis sample, but included in a sensitivity analysis that yielded largely similar results. It is possible that additional respondents who did not self-describe their gender completed the survey in bad faith.

A strength of our novel sampling method is that it generated a large sample with diverse characteristics enabling detailed subgroup analyses that identified new findings. For example, most previous studies of COVID-19 vaccine hesitancy grouped Asians with American Indian/Alaska Native, Native Hawaiian or other Pacific Islanders (13,40–42). However, our study, which included 11,962 Asian participants in the May sample, identified a remarkably lower prevalence of hesitancy among Asians versus all other race/ethnic groups. This study also identified an interaction between race/ethnicity and age (e.g., Blacks had relatively high hesitancy among adults under 35 years while Whites had relatively high hesitancy among adults 45 and older), which have previously been reported as independent predictors of vaccine hesitancy without investigating an interaction (13,43). Thus, our study provides more recent and extensive data to inform race/ethnicity and age orientated COVID-19 vaccination uptake efforts. Additionally, while a previous study evaluated changes in hesitancy by age, sex, education, or income level, October 2020 through March 2021 (13), estimated change by these categories had large overlapping 95% CI, and the racial and education categories collapsed groups in which we have identified meaningful differences.

## Conclusion

This massive national survey administered throughout the US COVID-19 vaccine rollout (January-May, 2021) identified a decrease in vaccine hesitancy by one-third, with important changes in hesitancy in key subgroups. This study also provided a fine-grained analysis of COVID-19 vaccine hesitancy by subgroup at the time of universal vaccine availability, and provided insight into reasons for hesitancy both overall and by key subgroups, revealing that somewhat hesitant and strongly hesitant adults indicate different beliefs about vaccine safety. These results can support the development of targeted public health campaigns and policies to increase COVID-19 vaccine uptake. Additionally, as new diseases continue to emerge (44), these findings can provide insight for the planning of future vaccine rollouts.

## Data Availability

To protect the confidentiality of survey respondents, access to survey microdata is restricted. Access is available to any academic or nonprofit researchers under a data use agreement. Information on how to request access can be found at https://cmu-delphi.github.io/delphi-epidata/symptom-survey/. Requests are reviewed by the Carnegie Mellon University Office of Sponsored Programs and Facebook Data for Good.

https://cmu-delphi.github.io/delphi-epidata/symptom-survey/coding.html

## Authors contributions

Dr. Reinhart and Mr. Rubinstein had full access to all of the data and take responsibility for the integrity of the data and the accuracy of the data analysis.

Obtained funding, supervised study and acquired data: AR

Developed study concept and design: WK, RM, AR

Performed statistical analysis: MR

Drafting of the manuscript: WK, RM

Interpreted data: WK, RM, AR, MR

Critically revised manuscript for important intellectual content: WK, RM, AR, MR

## Other acknowledgements

We would like to thank the Delphi Group at Carnegie Mellon University for input and support on the survey instrument. Wichada La Motte-Kerr, MPH, of Delphi contributed to the development and deployment of the survey and received compensation for her contributions to the study. We thank Sarah LaRocca, PhD and Katherine Morris, PhD of Facebook for contributions to the survey instrument design.

## Supporting Information

**sAppendix**. Carnegie Mellon University (CMU) Delphi Group’s COVID Trends and Impact Survey (CTIS): select questions and response sets.

**sTable 1**. Participant flow for Carnegie Mellon University (CMU) Delphi Group’s COVID Trends and Impact Survey (CTIS) by month (January-May, 2021)

**sTable 2**. Weighted demographics of the report sample by month. The distribution of responses is provided including and excluding missing responses for each item.

**sTable 3**. COVID-19 vaccine receipt and intent by month (January-May, 2021), among US adults

**sTable 4**. COVID-19 vaccine hesitancy by race/ethnicity, education level, US region and county Trump vote share in the 2020 presidential election, respectively, by month (January-May, 2021), among US adults

**sTable 5**. COVID-19 vaccine hesitancy in May 2021 by age groups, stratified by race/ethnicity^a^ among US adults

**sTable 6**. COVID-19 vaccine hesitancy in May 2021 by race/ethnicity^a^, stratified by age groups, among US adults

**sTable 7**. COVID-19 vaccine hesitancy in May 2021 by health condition and status^a^ among US adults

**sTable 8**. Sensitivity analysis: COVID-19 vaccine hesitancy in May 2021 by demographics among US adults, including those that self-described gender (N=529,658)

**sTable 9**. Sensitivity analysis: COVID-19 vaccine hesitancy in May 2021 political/by COVID-19 environment, health status, beliefs and behaviors among US adults, including those that self-described gender (N=529,658)

**sTable 10**. Reasons for COVID-19 vaccine hesitancy in May 21 by race/ethnicity^a^ among hesitant US adults (N=73362)^b^

