## Supplemental Material for "Time trends, factors associated with, and reasons for COVID-19 vaccine hesitancy in a massive online survey of US adults: January-May 2021"

**Supporting Information**

**sAppendix**. Carnegie Mellon University (CMU) Delphi Group's COVID Trends and Impact Survey (CTIS): select questions and response sets.

**sTable 1**. Participant flow for Carnegie Mellon University (CMU) Delphi Group's COVID Trends and Impact Survey (CTIS) by month (January-May, 2021)

**sTable 2.** Weighted demographics of the report sample by month. The distribution of responses is provided including and excluding missing responses for each item.

**sTable 3.** COVID-19 vaccine receipt and intent by month (January-May, 2021), among US adults

**sTable 4**. COVID-19 vaccine hesitancy by race/ethnicity, education level, US region and county Trump vote share in the 2020 presidential election, respectively, by month (January-May, 2021), among US adults

**sTable 5.** COVID-19 vaccine hesitancy in May 2021 by age groups, stratified by race/ethnicity[a] among US adults

**sTable 6.** COVID-19 vaccine hesitancy in May 2021 by race/ethnicity [a], stratified by age groups, among US adults

**sTable 7**. COVID-19 vaccine hesitancy in May 2021 by health condition and status[a] among US adults

**sTable 8.** Sensitivity analysis: COVID-19 vaccine hesitancy in May 2021 by demographics among US adults, including those that self-described gender (N=529,658)

**sTable 9.** Sensitivity analysis: COVID-19 vaccine hesitancy in May 2021 political/by COVID-19 environment, health status, beliefs and behaviors among US adults, including those that self-described gender (N=529,658)

**sTable 10**. Reasons for COVID-19 vaccine hesitancy in May 21 by race/ethnicity[a] among hesitant US adults (N=73362)[b]

**sAppendix**. Carnegie Mellon University (CMU) Delphi Group's COVID Trends and Impact Survey (CTIS): select questions and response sets.

Below are the survey items used in this study. Unless otherwise noted, the survey items remained unchanged during the study period. The full text of each survey instrument is available online: https://cmu-delphi.github.io/delphi-epidata/symptom-survey/coding.html

D1 What is your gender?

1. Male
2. Female
3. Non-binary
4. Prefer to self-describe _________________
5. Prefer not to answer

D2 What is your age?

1. 18-24 years
2. 25-34 years
3. 35-44 years
4. 45-54 years
5. 55-64 years
6. 65-74 years
7. 75 years or older

D6 Are you of Hispanic, Latino, or Spanish origin?

1. Yes
2. No, not of Hispanic, Latino, or Spanish origin

D7 What is your race? Please select all that apply.

1. American Indian or Alaska Native
2. Asian
3. Black or African American
4. Native Hawaiian or other Pacific Islander
5. White
6. Some other race

Continued next page

D8 What is the highest degree or level of school you have completed?

1. Less than high school
2. High school graduate or equivalent (GED)
3. Some college
4. 2 year degree
5. 4 year degree
6. Master’s degree
7. Professional degree (e.g. MD, JD, DVM)
8. Doctorate

D9 In the past 4 weeks, did you do any kind of work for pay?

1. Yes
2. No

D10 [Displayed if D9 = “Yes”] Was any of your work for pay in the last four weeks outside your home?

1. Yes
2. No

A3 What is your current ZIP code? _____________

B11 Have you **ever** tested positive for coronavirus (COVID-19)?

1. Yes
2. No
3. I don't know

C1 Have you ever been told by a doctor, nurse, or other health professional that you have any of the following medical conditions? Please select all that apply.

1. Cancer (other than skin cancer)
2. Heart attack, heart disease, or other heart condition
3. High blood pressure
4. Asthma
5. Chronic lung disease such as COPD, chronic bronchitis, or emphysema
6. Kidney disease
7. Autoimmune disorder such as rheumatoid arthritis or Crohn’s disease
8. Type 1 diabetes
9. Type 2 diabetes
10. Weakened or compromised immune system
11. Obesity
12. None of these

A5 How many people, including you, are currently staying in your household?

1. Children under 18 years old _____________
2. Adults between 18 and 64 years old _____________
3. Adults 65 years old or older ______________

C9 How worried do you feel that you or someone in your immediate family might become seriously ill from COVID-19 (coronavirus disease)?

1. Very worried
2. Somewhat worried
3. Not too worried
4. Not worried at all

C17 Have you gotten a seasonal flu vaccine since June 2020?

1. Yes, I have gotten a seasonal flu vaccine since June 2020
2. No, I have not gotten a seasonal flu vaccine since June 2020
3. I don't know if I have gotten a seasonal flu vaccine since June 2020

C7 To what extent are you intentionally avoiding contact with other people?

1. All of the time
2. Most of the time; I only leave my home to buy food and other essentials
3. Some of the time; I have reduced the amount of times I am in public spaces, social gatherings, or at work
4. None of the time

V1 Have you had a COVID-19 vaccination?

1. Yes
2. No
3. I don't know

V3 If a vaccine to prevent COVID-19 were offered to you today, would you choose to get vaccinated?

1. Yes, definitely
2. Yes, probably
3. No, probably not
4. No, definitely not

V5

a) [Displayed if V3 = “Yes, probably”] Which of the following, if any, are reasons that you only probably will* get a COVID-19 vaccine? Please select all that apply.
b) [Displayed if V3 = “No, probably not”] Which of the following, if any, are reasons that you probably won’t* get a COVID-19 vaccine? Please select all that apply.
c) [Displayed if V3 = “No, definitely not”] Which of the following, if any, are reasons that you definitely won’t* get a COVID-19 vaccine? Please select all that apply.

*The wording of the response sets was changed from “will get” to “would choose to get,” and “won’t get” to “wouldn’t choose to get,” on February 8, 2021.

Answer choices:

1. I am concerned about possible side effects of a COVID-19 vaccine.
2. I am concerned about having an allergic reaction to a COVID-19 vaccine.
3. I don't know if a COVID-19 vaccine will work.
4. I don't believe I need a COVID-19 vaccine.
5. I don't like vaccines.
6. My doctor has not recommended it.
7. I plan to wait and see if it is safe and may get it later.
8. I think other people need it more than I do right now.
9. I am concerned about the cost of a COVID-19 vaccine.
10. I don't trust COVID-19 vaccines.
11. I don't trust the government.
12. It is against my religious beliefs.
13. I have a health condition and am concerned about the safety of the vaccine for people with my condition.
14. I am currently/planning to be pregnant and/or breastfeeding and do not want to get vaccinated at this time.
15. Other

**sTable 1**. Participant flow for Carnegie Mellon University (CMU) Delphi Group's COVID Trends and Impact Survey (CTIS) by month (January-May, 2021)

| | **January[a]** | **February** | **March** | **April** | **May [b]** | **Total** |
|---|---|---|---|---|---|---|
| | **N** | | | | | |
| Offered survey | 106,387,320 | 95,953,902 | 104,768,154 | 103,399,752 | 66,138,989 | 476,648,117 |
| Did not respond | 105,073,194 | 94,721,211 | 103,476,197 | 102,315,236 | 65,576,417 | 471,162,255 |
| Responded | 1,314,126 | 1,232,691 | 1,291,957 | 1,084,516 | 562,572 | 5,485,862 |
| Response rate | 1.24% | 1.28% | 1.23% | 1.05% | 0.85% | 1.15% |
| Did not report hesitancy | 112,319 | 83,829 | 74468 | 61896 | 32,914 | 365,426 |
| Reported self-describe gender[c] | 6,157 | 6,667 | 7,953 | 6,873 | 4,014 | 31,664 |
| **Report sample** | 1,195,650 | 1,142,195 | 1,209,536 | 1,015,747 | 525,644 | 5,088,772 |
| Vaccinated/Definitely Yes | x | x | x | x | 435,393 | x |
| Did not provide reasons | x | x | x | x | 2,267 | x |
| **Reasons sample** | x | x | x | x | 87,984 | x |

[a] January data was subset to January 6-31 because January 6 was the first date both vaccine questions (receipt and intent) were both asked.

[b] May data was subset to include only those receiving the survey prior to an update to the vaccine questions; on May 20 a new survey version was offered to approximately 85% of potential respondents**.**

[c] A review of fill-in responses for self-described gender suggested the majority of participants who selected this category did not complete the survey in good faith (e.g., wrote-in discriminatory statements, and as a group had a high frequency of extreme responses such as the oldest age group and highest education level). Thus, they were excluded from the main analysis. They are included in a sensitivity analysis (sTable 8-9).

**sTable 2.** Weighted demographics of the report sample by month. The distribution of responses is provided including and excluding missing responses for each item.

| | **January** | | **February** | | **March** | | **April** | | **May** | |
|---|---|---|---|---|---|---|---|---|---|---|
| | Full sample | Not missing | Full sample | Not missing | Full sample | Not missing | Full sample | Not missing | Full sample | Not missing |
| **Gender** | | | | | | | | | | |
| Male | 40.04 | 46.03 | 39.34 | 46.09 | 38.79 | 46.01 | 38.93 | 45.94 | 38.76 | 45.68 |
| Female | 46.19 | 53.11 | 45.25 | 53.02 | 44.75 | 53.09 | 44.96 | 53.06 | 45.13 | 53.18 |
| Non-binary | 0.75 | 0.86 | 0.76 | 0.89 | 0.76 | 0.90 | 0.84 | 1.00 | 0.97 | 1.14 |
| Missing | 13.03 | NA | 14.65 | NA | 15.70 | NA | 15.26 | NA | 15.14 | NA |
| **Age group** | | | | | | | | | | |
| 18-24 years | 9.03 | 10.26 | 8.73 | 10.13 | 8.46 | 9.94 | 8.40 | 9.82 | 8.46 | 9.87 |
| 25-34 years | 14.16 | 16.08 | 13.64 | 15.84 | 13.28 | 15.60 | 13.37 | 15.62 | 13.56 | 15.82 |
| 35-44 years | 14.42 | 16.39 | 14.21 | 16.50 | 14.02 | 16.47 | 14.18 | 16.57 | 14.13 | 16.49 |
| 45-54 years | 15.40 | 17.50 | 14.98 | 17.39 | 14.72 | 17.29 | 14.88 | 17.39 | 14.72 | 17.17 |
| 55-64 years | 15.65 | 17.79 | 15.41 | 17.89 | 15.43 | 18.12 | 15.64 | 18.28 | 15.81 | 18.45 |
| 65-74 years | 13.48 | 15.31 | 13.34 | 15.48 | 13.49 | 15.85 | 13.40 | 15.66 | 13.24 | 15.46 |
| ≥ 75 years | 5.87 | 6.67 | 5.83 | 6.77 | 5.72 | 6.72 | 5.70 | 6.66 | 5.78 | 6.75 |
| Missing | 11.99 | NA | 13.86 | NA | 14.88 | NA | 14.44 | NA | 14.31 | NA |
| **Race/ethnicity** | | | | | | | | | | |
| White | 59.70 | 69.21 | 58.49 | 69.39 | 57.63 | 69.16 | 57.84 | 69.08 | 57.87 | 69.07 |
| Hispanic | 14.68 | 17.02 | 14.14 | 16.78 | 13.82 | 16.59 | 13.90 | 16.60 | 13.77 | 16.44 |
| Black | 6.33 | 7.34 | 6.10 | 7.23 | 5.76 | 6.91 | 5.54 | 6.62 | 5.46 | 6.52 |
| Asian | 2.42 | 2.80 | 2.46 | 2.92 | 2.51 | 3.01 | 2.84 | 3.39 | 3.01 | 3.59 |
| Native American | 0.92 | 1.07 | 0.89 | 1.05 | 0.75 | 0.90 | 0.71 | 0.84 | 0.71 | 0.84 |
| Pacific Islander | 0.23 | 0.26 | 0.23 | 0.27 | 0.21 | 0.26 | 0.21 | 0.25 | 0.21 | 0.24 |
| Multi-racial | 1.98 | 2.30 | 1.99 | 2.36 | 2.65 | 3.17 | 2.69 | 3.22 | 2.76 | 3.30 |
| Unknown (other or missing) | 13.74 | NA | 15.71 | NA | 16.67 | NA | 16.28 | NA | 16.22 | NA |

Continued on next page

| | | | | | | | | | | |
|---|---|---|---|---|---|---|---|---|---|---|
| **Education level** | | | | | | | | | | |
| ≤ High school | 20.91 | 24.05 | 19.94 | 23.43 | 19.01 | 22.61 | 18.90 | 22.38 | 19.03 | 22.51 |
| Some college | 32.00 | 36.81 | 31.13 | 36.59 | 30.73 | 36.55 | 30.68 | 36.33 | 30.84 | 36.46 |
| 4 year degree | 19.76 | 22.73 | 19.57 | 23.00 | 19.83 | 23.58 | 20.20 | 23.91 | 20.12 | 23.79 |
| Master's | 10.15 | 11.67 | 10.27 | 12.07 | 10.35 | 12.30 | 10.44 | 12.36 | 10.30 | 12.18 |
| Professional (e.g., MD, JD) | 2.50 | 2.87 | 2.53 | 2.98 | 2.53 | 3.01 | 2.54 | 3.01 | 2.57 | 3.03 |
| Doctorate | 1.61 | 1.86 | 1.64 | 1.93 | 1.63 | 1.94 | 1.70 | 2.01 | 1.71 | 2.02 |
| No response | 13.06 | NA | 14.92 | NA | 15.91 | NA | 15.54 | NA | 15.43 | NA |
| **Employment status** | | | | | | | | | | |
| Work outside home | 35.66 | 41.44 | 35.27 | 41.80 | 35.39 | 42.41 | 35.59 | 42.48 | 35.99 | 42.89 |
| Work at home | 11.81 | 13.72 | 11.06 | 13.11 | 10.82 | 12.97 | 10.95 | 13.07 | 10.61 | 12.65 |
| Does not work for pay | 38.59 | 44.84 | 38.04 | 45.09 | 37.23 | 44.62 | 37.24 | 44.45 | 37.31 | 44.46 |
| Missing | 13.95 | NA | 15.63 | NA | 16.55 | NA | 16.21 | NA | 16.09 | NA |

**sTable 3.** COVID-19 vaccine receipt and intent by month (January-May, 2021), among US adults

| | January | February | March | April | May | Difference (May - January) |
|---|---|---|---|---|---|---|
| Already vaccinated | 10.9 (10.8, 10.9) | 24.3 (24.2, 24.4) | 44.7 (44.6, 44.8) | 68.3 (68.1, 68.4) | 76.4 (76.2, 76.6) | 65.5 (65.4, 65.7) |
| Yes, definitely | 45.4 (45.3, 45.5) | 37.4 (37.3, 37.5) | 24.3 (24.2, 24.4) | 8.1 (8.0, 8.2) | 3.3 (3.2, 3.4) | -42.1 (-42.2, -42.0) |
| Yes, probably | 18.3 (18.2, 18.4) | 14.6 (14.5, 14.7) | 10.6 (10.5, 10.7) | 5.9 (5.8, 6.0) | 3.7 (3.7, 3.8) | -14.6 (-14.7, -14.5) |
| No, probably not | 13.0 (13.0, 13.1) | 11.5 (11.5, 11.6) | 9.3 (9.2, 9.3) | 7.1 (7.0, 7.1) | 6.0 (5.9, 6.1) | -7.1 (-7.2, -6.9) |
| No, definitely not | 12.4 (12.3, 12.4) | 12.1 (12.1, 12.2) | 11.0 (11.0, 11.1) | 10.7 (10.6, 10.7) | 10.6 (10.5, 10.7) | -1.8 (-1.9, -1.7) |
| Hesitant[a] | 25.4 (25.3, 25.5) | 23.7 (23.6, 23.8) | 20.3 (20.2, 20.4) | 17.7 (17.6, 17.8) | 16.6 (16.4, 16.7) | -8.9 (-9.0, -8.7) |

[a] Answered that they probably or definitely would not choose to get vaccinated if offered a vaccine to prevent COVID-19

**sTable 4**. COVID-19 vaccine hesitancy by race/ethnicity, education level, US region and county Trump vote share in the 2020 presidential election, respectively, by month (January-May, 2021), among US adults

| | January | February | March | April | May | Difference (May - January) |
|---|---|---|---|---|---|---|
| | % (95% CI) | | | | | |
| Race/ethnicity among 18 - 34 year olds[a] | | | | | | |
| White | 31.0 (30.7, 31.3) | 29.7 (29.4, 30.0) | 27.2 (26.8, 27.5) | 23.9 (23.6, 24.3) | 22.5 (22.0, 23.0) | -8.5 (-9.1, -7.9) |
| Hispanic | 31.7 (31.2, 32.3) | 28.5 (27.9, 29.1) | 23.6 (23.0, 24.2) | 19.0 (18.4, 19.7) | 16.3 (15.5, 17.1) | -15.4 (-16.4, -14.4) |
| Black | 58.9 (57.7, 60.0) | 53.0 (51.8, 54.3) | 43.1 (41.8, 44.5) | 34.0 (32.5, 35.6) | 27.4 (25.3, 29.4) | -31.5 (-33.8, -29.2) |
| Asian | 12.4 (11.4, 13.4) | 11.6 (10.6, 12.7) | 8.0 (7.1, 8.9) | 4.6 (3.9, 5.2) | 3.2 (2.4, 4.0) | -9.2 (-10.5, -7.9) |
| Native American | 43.2 (40.4, 46.0) | 39.4 (36.4, 42.4) | 32.4 (29.2, 35.7) | 30.8 (27.3, 34.4) | 33.7 (28.1, 39.3) | -9.4 (-15.7, -3.2) |
| Pacific Islander | 43.0 (37.0, 49.0) | 30.5 (24.9, 36.1) | 30.8 (24.3, 37.4) | 27.0 (18.4, 35.5) | 15.8 (7.9, 23.6) | -27.2 (-37.1, -17.4) |
| Multi-racial | 38.2 (36.9, 39.6) | 37.6 (36.2, 39.1) | 35.5 (34.1, 36.8) | 31.2 (29.7, 32.7) | 29.0 (26.9, 31.1) | -9.2 (-11.7, -6.7) |
| Education level | | | | | | |
| ≤ High school | 34.8 (34.6, 35.1) | 31.9 (31.6, 32.2) | 26.5 (26.2, 26.7) | 22.9 (22.6, 23.2) | 20.3 (20.0, 20.7) | -14.5 (-14.9, -14.0) |
| Some college | 27.2 (27.0, 27.3) | 25.3 (25.1, 25.5) | 21.6 (21.5, 21.8) | 18.9 (18.7, 19.1) | 17.9 (17.7, 18.2) | -9.2 (-9.5, -9.0) |
| 4 year degree | 15.2 (15.1, 15.4) | 14.3 (14.2, 14.5) | 12.2 (12.0, 12.4) | 10.8 (10.7, 11.0) | 10.8 (10.5, 11.0) | -4.5 (-4.8, -4.2) |
| Master's | 11.5 (11.4, 11.7) | 10.5 (10.3, 10.7) | 9.0 (8.8, 9.2) | 8.0 (7.8, 8.2) | 7.8 (7.6, 8.1) | -3.7 (-4.0, -3.4) |
| Professional (e.g., MD, JD) | 11.9 (11.4, 12.3) | 11.8 (11.3, 12.2) | 11.0 (10.6, 11.4) | 10.0 (9.5, 10.4) | 11.1 (10.4, 11.8) | -0.8 (-1.6, 0.0) |
| Doctorate | 16.5 (15.8, 17.1) | 16.1 (15.4, 16.8) | 14.6 (13.9, 15.2) | 14.8 (14.1, 15.5) | 14.6 (13.5, 15.6) | -1.9 (-3.1, -0.7) |
| US Region | | | | | | |
| Midwest | 26.3 (26.1, 26.5) | 24.7 (24.5, 24.9) | 21.6 (21.5, 21.8) | 19.0 (18.8, 19.2) | 17.7 (17.4, 17.9) | -8.6 (-9.0, -8.3) |
| South | 28.5 (28.3, 28.7) | 26.8 (26.6, 26.9) | 22.9 (22.7, 23.1) | 20.0 (19.8, 20.2) | 18.8 (18.6, 19.1) | -9.7 (-10.0, -9.4) |
| Pacific | 20.0 (19.7, 20.2) | 18.0 (17.8, 18.3) | 15.2 (15.0, 15.4) | 12.6 (12.3, 12.8) | 11.7 (11.3, 12.0) | -8.3 (-8.7, -7.9) |
| Mountain | 24.4 (24.0, 24.7) | 22.9 (22.6, 23.3) | 20.3 (19.9, 20.6) | 18.2 (17.9, 18.6) | 17.5 (17.0, 18.0) | -6.8 (-7.4, -6.2) |
| Northeast | 21.6 (21.4, 21.8) | 19.6 (19.4, 19.8) | 16.1 (15.8, 16.3) | 13.7 (13.5, 13.9) | 12.2 (11.9, 12.5) | -9.4 (-9.7, -9.0) |
| County Trump vote share | | | | | | |
| Lowest quartile | 21.7 (21.6, 21.8) | 19.7 (19.6, 19.8) | 16.3 (16.2, 16.4) | 13.7 (13.6, 13.8) | 12.4 (12.3, 12.6) | -9.3 (-9.5, -9.1) |
| Second lowest quartile | 29.7 (29.5, 29.9) | 28.4 (28.2, 28.7) | 25.2 (24.9, 25.4) | 22.5 (22.3, 22.7) | 21.4 (21.1, 21.8) | -8.3 (-8.7, -7.9) |
| Second highest quartile | 35.1 (34.7, 35.4) | 33.6 (33.3, 34.0) | 30.2 (29.9, 30.5) | 27.9 (27.5, 28.3) | 27.0 (26.5, 27.6) | -8.1 (-8.7, -7.4) |
| Highest quartile | 38.8 (38.2, 39.3) | 38.4 (37.8, 38.9) | 35.4 (34.8, 35.9) | 32.6 (32.0, 33.2) | 31.9 (31.1, 32.8) | -6.8 (-7.8, -5.8) |

Juris Doctorate= JD; MD=Doctor of Medicine.
[a] Race/ethnicity categories are reported among young adults (18-34 years) only, because there was an interaction between race/ethnicity and age, such that race/ethnic comparisons differed by age group. Younger adults were selected because hesitancy was higher among younger versus older adults. Race/ethnicity groups other than the group labeled “Hispanic” are non-Hispanic.

**sTable 5.** COVID-19 vaccine hesitancy in May 2021 by age groups, stratified by race/ethnicity[a] among US adults

| | Sample | | COVID-19 vaccine hesitant | | |
|---|---|---|---|---|---|
| | **n** | **%** | **% (95% CI)** | **RR (95% CI)** | **Adj. RR (95% CI)** |
| White | | | | | |
| 18-24 years | 9858 | 1.9 | 23.5 (22.5, 24.6) | 2.89 (2.75, 3.04) | 1.46 (1.38, 1.53) |
| 25-34 years | 35244 | 6.7 | 21.9 (21.3, 22.4) | 2.69 (2.60, 2.79) | 1.64 (1.57, 1.70) |
| 35-44 years | 49337 | 9.4 | 18.6 (18.2, 19.0) | 2.29 (2.21, 2.37) | 1.47 (1.42, 1.53) |
| 45-54 years | 54805 | 10.4 | 18.2 (17.9, 18.6) | 2.24 (2.17, 2.32) | 1.42 (1.37, 1.48) |
| 55-64 years | 74941 | 14.3 | 13.3 (13.0, 13.6) | 1.64 (1.58, 1.69) | 1.21 (1.16, 1.25) |
| 65-74 years | 76794 | 14.6 | 8.1 (7.9, 8.3) | 1.00 (NA) | 1.00 (NA) |
| ≥ 75 years | 36019 | 6.9 | 6.0 (5.7, 6.2) | 0.73 (0.69, 0.77) | 0.76 (0.73, 0.80) |
| Hispanic | | | | | |
| 18-24 years | 3043 | 0.6 | 17.1 (15.5, 18.7) | 2.84 (2.44, 3.24) | 1.64 (1.42, 1.86) |
| 25-34 years | 8668 | 1.6 | 15.7 (14.8, 16.6) | 2.61 (2.30, 2.92) | 1.74 (1.54, 1.93) |
| 35-44 years | 11700 | 2.2 | 12.5 (11.7, 13.3) | 2.08 (1.82, 2.34) | 1.42 (1.25, 1.59) |
| 45-54 years | 12943 | 2.5 | 9.0 (8.5, 9.6) | 1.50 (1.32, 1.69) | 1.13 (1.00, 1.26) |
| 55-64 years | 11507 | 2.2 | 7.7 (7.1, 8.2) | 1.27 (1.11, 1.44) | 1.05 (0.93, 1.18) |
| 65-74 years | 6481 | 1.2 | 6.0 (5.4, 6.7) | 1.00 (NA) | 1.00 (NA) |
| ≥ 75 years | 1921 | 0.4 | 15.2 (13.1, 17.4) | 2.53 (2.08, 2.97) | 1.61 (1.39, 1.84) |
| Black | | | | | |
| 18-24 years | 626 | 0.1 | 30.9 (26.6, 35.2) | 7.26 (5.92, 8.61) | 3.51 (2.77, 4.24) |
| 25-34 years | 2205 | 0.4 | 25.3 (23.3, 27.4) | 5.96 (5.09, 6.83) | 3.73 (3.18, 4.28) |
| 35-44 years | 3785 | 0.7 | 18.7 (17.3, 20.0) | 4.39 (3.77, 5.01) | 2.96 (2.54, 3.37) |
| 45-54 years | 5536 | 1.1 | 12.2 (11.2, 13.1) | 2.86 (2.45, 3.28) | 2.13 (1.83, 2.44) |
| 55-64 years | 8289 | 1.6 | 7.8 (7.2, 8.5) | 1.84 (1.57, 2.11) | 1.47 (1.26, 1.69) |
| 65-74 years | 6390 | 1.2 | 4.3 (3.7, 4.8) | 1.00 (NA) | 1.00 (NA) |
| ≥ 75 years | 1669 | 0.3 | 3.0 (2.1, 4.0) | 0.71 (0.47, 0.95) | 0.75 (0.51, 0.98) |
| Asian | | | | | |
| 18-24 years | 612 | 0.1 | 4.0 (2.1, 6.0) | 2.45 (0.96, 3.94) | 1.68 (0.68, 2.68) |
| 25-34 years | 2288 | 0.4 | 2.7 (2.0, 3.4) | 1.65 (0.90, 2.41) | 1.42 (0.78, 2.06) |
| 35-44 years | 2745 | 0.5 | 2.9 (2.2, 3.6) | 1.75 (0.96, 2.55) | 1.56 (0.87, 2.24) |
| 45-54 years | 2234 | 0.4 | 3.3 (2.4, 4.2) | 2.02 (1.08, 2.95) | 1.70 (0.93, 2.47) |
| 55-64 years | 1957 | 0.4 | 2.8 (2.0, 3.6) | 1.69 (0.88, 2.49) | 1.50 (0.80, 2.21) |
| 65-74 years | 1527 | 0.3 | 1.7 (1.0, 2.3) | 1.00 (NA) | 1.00 (NA) |
| ≥ 75 years | 586 | 0.1 | 2.4 (0.9, 3.9) | 1.48 (0.41, 2.54) | 1.51 (0.44, 2.58) |
| Native American | | | | | |
| 18-24 years | 142 | 0.0 | 37.3 (26.0, 48.7) | 2.76 (1.73, 3.79) | 1.60 (0.95, 2.24) |
| 25-34 years | 406 | 0.1 | 31.3 (25.8, 36.7) | 2.31 (1.66, 2.96) | 1.28 (0.98, 1.58) |
| 35-44 years | 574 | 0.1 | 27.2 (23.1, 31.4) | 2.01 (1.48, 2.55) | 1.32 (1.03, 1.61) |
| 45-54 years | 841 | 0.2 | 24.3 (21.1, 27.6) | 1.80 (1.34, 2.26) | 1.19 (0.94, 1.45) |
| 55-64 years | 1031 | 0.2 | 21.8 (19.0, 24.6) | 1.61 (1.20, 2.02) | 1.18 (0.93, 1.43) |
| 65-74 years | 691 | 0.1 | 13.5 (10.6, 16.5) | 1.00 (NA) | 1.00 (NA) |
| ≥ 75 years | 244 | 0.0 | 17.7 (12.1, 23.3) | 1.31 (0.80, 1.81) | 1.36 (0.92, 1.81) |

Continued next page

| | | | | | |
|---|---|---|---|---|---|
| Pacific Islander[a] | | | | | |
| 18-24 years | 20 | 0.0 | 6.0 (-4.3, 16.2) | 0.81 (-0.66, 2.28) | 0.62 (-0.45, 1.69) |
| 25-34 years | 68 | 0.0 | 21.2 (11.2, 31.1) | 2.87 (0.77, 4.97) | 1.47 (0.46, 2.47) |
| 35-44 years | 150 | 0.0 | 14.1 (7.9, 20.4) | 1.92 (0.55, 3.29) | 1.46 (0.50, 2.43) |
| 45-54 years | 230 | 0.0 | 16.5 (10.8, 22.1) | 2.24 (0.77, 3.70) | 1.54 (0.59, 2.48) |
| 55-64 years | 277 | 0.1 | 11.6 (7.2, 16.1) | 1.58 (0.51, 2.65) | 1.28 (0.46, 2.09) |
| 65-74 years | 190 | 0.0 | 7.4 (3.3, 11.5) | 1.00 (NA) | 1.00 (NA) |
| ≥ 75 years | 56 | 0.0 | 14.5 (2.6, 26.4) | 1.97 (0.01, 3.92) | 1.64 (0.44, 2.83) |
| Multi-racial | | | | | |
| 18-24 years | 880 | 0.2 | 29.7 (26.0, 33.4) | 1.90 (1.58, 2.23) | 1.20 (1.03, 1.38) |
| 25-34 years | 2209 | 0.4 | 28.5 (26.3, 30.7) | 1.83 (1.57, 2.08) | 1.27 (1.11, 1.42) |
| 35-44 years | 2557 | 0.5 | 30.0 (28.0, 32.0) | 1.92 (1.67, 2.18) | 1.29 (1.13, 1.44) |
| 45-54 years | 2385 | 0.5 | 28.3 (26.3, 30.4) | 1.82 (1.57, 2.07) | 1.23 (1.08, 1.38) |
| 55-64 years | 2491 | 0.5 | 24.6 (22.7, 26.5) | 1.58 (1.36, 1.80) | 1.16 (1.02, 1.30) |
| 65-74 years | 1803 | 0.3 | 15.6 (13.8, 17.4) | 1.00 (NA) | 1.00 (NA) |
| ≥ 75 years | 660 | 0.1 | 17.6 (13.6, 21.6) | 1.13 (0.84, 1.42) | 0.89 (0.72, 1.05) |

[a] Race/ethnicity groups other than the group labeled “Hispanic” are non-Hispanic.

[b] The Hispanic ≥ 75 years group has unusually high rates of self-describe gender; results were attenuated in a sensitivity analysis with that group (self-describe gender) removed.

[c] Due to the limited sample size of age groups among Pacific Islanders, the 95% CI of the RR estimates are large.

**sTable 6.** COVID-19 vaccine hesitancy in May 2021 by race/ethnicity [a], stratified by age groups, among US adults

| | Sample | | COVID-19 vaccine hesitant | | |
|---|---|---|---|---|---|
| | **n** | **%** | **% (95% CI)** | **RR (95% CI)** | **Adj. RR (95% CI)** |
| 18-24 years | | | | | |
| White | 9858 | 1.9 | 23.5 (22.5, 24.6) | 1.00 (NA) | 1.00 (NA) |
| Hispanic | 3043 | 0.6 | 17.1 (15.5, 18.7) | 0.73 (0.65, 0.80) | 0.92 (0.84, 1.01) |
| Black | 626 | 0.1 | 30.9 (26.6, 35.2) | 1.31 (1.12, 1.51) | 1.47 (1.21, 1.73) |
| Asian | 612 | 0.1 | 4.0 (2.1, 6.0) | 0.17 (0.09, 0.25) | 0.38 (0.20, 0.55) |
| Native American | 142 | 0.0 | 37.3 (26.0, 48.7) | 1.59 (1.10, 2.08) | 1.43 (0.92, 1.95) |
| Pacific Islander | 20 | 0.0 | 6.0 (-4.3, 16.2) | 0.25 (-0.18, 0.69) | 0.44 (-0.29, 1.17) |
| Multi-racial | 880 | 0.2 | 29.7 (26.0, 33.4) | 1.26 (1.09, 1.43) | 1.26 (1.12, 1.39) |
| 25-34 years | | | | | |
| White | 35244 | 6.7 | 21.9 (21.3, 22.4) | 1.00 (NA) | 1.00 (NA) |
| Hispanic | 8668 | 1.6 | 15.7 (14.8, 16.6) | 0.72 (0.68, 0.76) | 0.87 (0.82, 0.92) |
| Black | 2205 | 0.4 | 25.3 (23.3, 27.4) | 1.16 (1.06, 1.26) | 1.38 (1.26, 1.50) |
| Asian | 2288 | 0.4 | 2.7 (2.0, 3.4) | 0.12 (0.09, 0.16) | 0.28 (0.21, 0.36) |
| Native American | 406 | 0.1 | 31.3 (25.8, 36.7) | 1.43 (1.18, 1.68) | 1.02 (0.87, 1.18) |
| Pacific Islander | 68 | 0.0 | 21.2 (11.2, 31.1) | 0.97 (0.51, 1.43) | 0.93 (0.52, 1.35) |
| Multi-racial | 2209 | 0.4 | 28.5 (26.3, 30.7) | 1.30 (1.20, 1.41) | 1.18 (1.09, 1.26) |
| 35-44 years | | | | | |
| White | 49337 | 9.4 | 18.6 (18.2, 19.0) | 1.00 (NA) | 1.00 (NA) |
| Hispanic | 11700 | 2.2 | 12.5 (11.7, 13.3) | 0.67 (0.63, 0.72) | 0.79 (0.74, 0.84) |
| Black | 3785 | 0.7 | 18.7 (17.3, 20.0) | 1.00 (0.93, 1.08) | 1.22 (1.13, 1.32) |
| Asian | 2745 | 0.5 | 2.9 (2.2, 3.6) | 0.16 (0.12, 0.20) | 0.34 (0.26, 0.43) |
| Native American | 574 | 0.1 | 27.2 (23.1, 31.4) | 1.46 (1.24, 1.69) | 1.17 (1.03, 1.32) |
| Pacific Islander | 150 | 0.0 | 14.1 (7.9, 20.4) | 0.76 (0.42, 1.10) | 1.04 (0.62, 1.46) |
| Multi-racial | 2557 | 0.5 | 30.0 (28.0, 32.0) | 1.61 (1.50, 1.73) | 1.33 (1.25, 1.41) |
| 45-54 years | | | | | |
| White | 54805 | 10.4 | 18.2 (17.9, 18.6) | 1.00 (NA) | 1.00 (NA) |
| Hispanic | 12943 | 2.5 | 9.0 (8.5, 9.6) | 0.50 (0.47, 0.53) | 0.65 (0.61, 0.69) |
| Black | 5536 | 1.1 | 12.2 (11.2, 13.1) | 0.67 (0.61, 0.72) | 0.91 (0.84, 0.98) |
| Asian | 2234 | 0.4 | 3.3 (2.4, 4.2) | 0.18 (0.13, 0.23) | 0.39 (0.29, 0.49) |
| Native American | 841 | 0.2 | 24.3 (21.1, 27.6) | 1.33 (1.16, 1.51) | 1.10 (0.97, 1.23) |
| Pacific Islander | 230 | 0.0 | 16.5 (10.8, 22.1) | 0.90 (0.59, 1.21) | 1.13 (0.76, 1.49) |
| Multi-racial | 2385 | 0.5 | 28.3 (26.3, 30.4) | 1.55 (1.44, 1.67) | 1.32 (1.23, 1.40) |
| 55-64 years | | | | | |
| White | 74941 | 14.3 | 13.3 (13.0, 13.6) | 1.00 (NA) | 1.00 (NA) |
| Hispanic | 11507 | 2.2 | 7.7 (7.1, 8.2) | 0.58 (0.53, 0.62) | 0.71 (0.66, 0.76) |
| Black | 8289 | 1.6 | 7.8 (7.2, 8.5) | 0.59 (0.54, 0.64) | 0.74 (0.68, 0.81) |
| Asian | 1957 | 0.4 | 2.8 (2.0, 3.6) | 0.21 (0.15, 0.27) | 0.41 (0.29, 0.52) |
| Native American | 1031 | 0.2 | 21.8 (19.0, 24.6) | 1.64 (1.42, 1.85) | 1.27 (1.14, 1.41) |
| Pacific Islander | 277 | 0.1 | 11.6 (7.2, 16.1) | 0.88 (0.54, 1.21) | 1.10 (0.69, 1.51) |
| Multi-racial | 2491 | 0.5 | 24.6 (22.7, 26.5) | 1.85 (1.71, 2.00) | 1.46 (1.37, 1.56) |

Continued next page

| | | | | | |
|---|---|---|---|---|---|
| 65-74 years | | | | | |
| White | 76794 | 14.6 | 8.1 (7.9, 8.3) | 1.00 (NA) | 1.00 (NA) |
| Hispanic | 6481 | 1.2 | 6.0 (5.4, 6.7) | 0.74 (0.66, 0.82) | 0.82 (0.74, 0.90) |
| Black | 6390 | 1.2 | 4.3 (3.7, 4.8) | 0.52 (0.46, 0.59) | 0.61 (0.53, 0.68) |
| Asian | 1527 | 0.3 | 1.7 (1.0, 2.3) | 0.20 (0.13, 0.28) | 0.33 (0.21, 0.45) |
| Native American | 691 | 0.1 | 13.5 (10.6, 16.5) | 1.67 (1.30, 2.03) | 1.31 (1.07, 1.55) |
| Pacific Islander | 190 | 0.0 | 7.4 (3.3, 11.5) | 0.91 (0.40, 1.41) | 1.04 (0.50, 1.59) |
| Multi-racial | 1803 | 0.3 | 15.6 (13.8, 17.4) | 1.92 (1.69, 2.15) | 1.52 (1.36, 1.68) |
| ≥ 75 years | | | | | |
| White | 36019 | 6.9 | 6.0 (5.7, 6.2) | 1.00 (NA) | 1.00 (NA) |
| Hispanic | 1921 | 0.4 | 15.2 (13.1, 17.4) | 2.56 (2.18, 2.94) | 1.73 (1.54, 1.92) |
| Black | 1669 | 0.3 | 3.0 (2.1, 4.0) | 0.50 (0.34, 0.67) | 0.59 (0.42, 0.77) |
| Asian | 586 | 0.1 | 2.4 (0.9, 3.9) | 0.41 (0.16, 0.66) | 0.65 (0.26, 1.03) |
| Native American | 244 | 0.0 | 17.7 (12.1, 23.3) | 2.97 (2.02, 3.91) | 2.33 (1.69, 2.98) |
| Pacific Islander | 56 | 0.0 | 14.5 (2.6, 26.4) | 2.43 (0.43, 4.43) | 2.23 (1.09, 3.38) |
| Multi-racial | 660 | 0.1 | 17.6 (13.6, 21.6) | 2.95 (2.27, 3.64) | 1.77 (1.48, 2.06) |

[a] Race/ethnicity groups other than the group labeled "Hispanic" are non-Hispanic.

[b] The Hispanic ≥ 75 years has unusually high rates of self-describe gender; results were attenuated in a sensitivity analysis with that group (self-describe gender) removed

**sTable 7**. COVID-19 vaccine hesitancy in May 2021 by health condition and status[a] among US adults

| | Sample | | COVID-19 vaccine hesitant | | |
|---|---|---|---|---|---|
| | **n** | **%** | **% (95% CI)** | **RR (95% CI)** | **Adj. RR (95% CI)** |
| None | 182782 | 34.8 | 18.7 (18.5, 19.0) | 1.0 (NA) | 1.0 (NA) |
| Cancer (other than skin) | 10355 | 2.0 | 8.9 (8.2, 9.5) | 0.47 (0.44, 0.51) | 0.80 (0.75, 0.85) |
| Diabetes Type II | 19671 | 3.7 | 9.6 (9.1, 10.1) | 0.51 (0.49, 0.54) | 0.83 (0.79, 0.86) |
| Obesity | 50985 | 9.7 | 10.9 (10.5, 11.3) | 0.58 (0.56, 0.60) | 0.79 (0.76, 0.81) |
| Diabetes Type I | 2614 | 0.5 | 12.2 (10.7, 13.8) | 0.65 (0.57, 0.74) | 0.86 (0.75, 0.96) |
| High blood pressure | 48953 | 9.3 | 13.2 (12.8, 13.6) | 0.70 (0.68, 0.73) | 0.95 (0.92, 0.98) |
| Kidney disease | 2344 | 0.4 | 13.3 (11.6, 15.0) | 0.71 (0.62, 0.80) | 1.07 (0.94, 1.19) |
| Chronic obstructive pulmonary disease | 6380 | 1.2 | 13.7 (12.5, 15.0) | 0.73 (0.67, 0.80) | 1.03 (0.96, 1.11) |
| Multiple conditions | 127693 | 24.3 | 14.0 (13.7, 14.2) | 0.75 (0.73, 0.76) | 1.13 (1.11, 1.15) |
| Heart attack, heart disease, or other heart condition | 12821 | 2.4 | 14.6 (13.9, 15.4) | 0.78 (0.74, 0.83) | 1.11 (1.06, 1.17) |
| Autoimmune disorder | 9700 | 1.8 | 15.1 (14.2, 16.0) | 0.80 (0.76, 0.85) | 1.06 (1.01, 1.12) |
| Asthma | 26313 | 5.0 | 16.6 (15.9, 17.2) | 0.89 (0.85, 0.92) | 1.03 (0.99, 1.06) |
| Weakened or compromised immune system | 4359 | 0.8 | 20.7 (19.1, 22.2) | 1.10 (1.02, 1.19) | 1.43 (1.33, 1.52) |
| No response | 20674 | 3.9 | 35.5 (34.6, 36.3) | 1.89 (1.84, 1.95) | 1.71 (1.66, 1.76) |

[a] Participants were categorized as having high blood pressure only, each of the other conditions with or without high blood pressure, or "multiple conditions," defined as at least two conditions excluding high blood pressure, which was relatively common and has limited support as a risk-factor for poor COVID-19 outcomes.

**sTable 8.** Sensitivity analysis: COVID-19 vaccine hesitancy in May 2021 by demographics among US adults, including those that self-described gender (N=529,658)

| | Sample | | COVID-19 vaccine hesitant | | |
|---|---|---|---|---|---|
| | **n** | **%** | **% (95% CI)** | **RR (95% CI)** | **Adj. RR (95% CI)** |
| Gender | | | | | |
| Male | 159427 | 30.1 | 16.6 (16.4, 16.9) | 1.0 (NA) | 1.0 (NA) |
| Female | 294983 | 55.7 | 13.2 (13.1, 13.4) | 0.79 (0.78, 0.81) | 1.12 (1.10, 1.14) |
| Non-binary | 3232 | 0.6 | 18.2 (16.1, 20.3) | 1.10 (0.97, 1.22) | 0.99 (0.88, 1.10) |
| Self-described | 4014 | 0.8 | 64.2 (62.3, 66.1) | 3.86 (3.73, 3.99) | 1.42 (1.37, 1.47) |
| Missing | 68002 | 12.8 | 26.3 (25.8, 26.7) | 1.58 (1.54, 1.61) | 1.39 (1.34, 1.44) |
| Age group | | | | | |
| 18-24 years | 15678 | 3.0 | 22.9 (22.1, 23.7) | 2.79 (2.67, 2.91) | [b] |
| 25-34 years | 52640 | 9.9 | 21.3 (20.8, 21.7) | 2.60 (2.52, 2.68) | |
| 35-44 years | 73245 | 13.8 | 18.4 (18.1, 18.8) | 2.25 (2.18, 2.32) | |
| 45-54 years | 81578 | 15.4 | 17.0 (16.7, 17.3) | 2.07 (2.01, 2.13) | |
| 55-64 years | 103380 | 19.5 | 12.9 (12.7, 13.1) | 1.57 (1.53, 1.62) | |
| 65-74 years | 95964 | 18.1 | 8.2 (8.0, 8.4) | 1.0 (NA) | |
| ≥ 75 years | 42657 | 8.1 | 9.8 (9.4, 10.2) | 1.20 (1.14, 1.25) | |
| Missing | 64516 | 12.2 | 24.6 (24.1, 25.0) | 3.00 (2.91, 3.09) | |
| Race/ethnicity | | | | | |
| White | 338578 | 63.9 | 15.8 (15.6, 16.0) | 1.0 (NA) | [b] |
| Hispanic | 57608 | 10.9 | 13.4 (13.0, 13.7) | 0.85 (0.82, 0.87) | |
| Black | 28625 | 5.4 | 13.0 (12.5, 13.5) | 0.82 (0.79, 0.86) | |
| Asian | 12012 | 2.3 | 3.2 (2.8, 3.6) | 0.20 (0.17, 0.23) | |
| Native American | 3993 | 0.8 | 25.3 (23.4, 27.2) | 1.60 (1.48, 1.72) | |
| Pacific Islander | 1002 | 0.2 | 13.9 (11.3, 16.5) | 0.88 (0.71, 1.04) | |
| Multi-racial | 13433 | 2.5 | 29.2 (28.2, 30.2) | 1.85 (1.78, 1.92) | |
| Missing | 74407 | 14.0 | 26.5 (26.1, 27.0) | 1.68 (1.65, 1.71) | |
| Education level | | | | | |
| ≤ High school | 92557 | 17.5 | 20.8 (20.4, 21.1) | 1.88 (1.83, 1.93) | 1.56 (1.52, 1.60) |
| Some college | 167096 | 31.5 | 18.3 (18.1, 18.6) | 1.66 (1.62, 1.70) | 1.37 (1.34, 1.40) |
| 4 year degree | 110944 | 20.9 | 11.0 (10.8, 11.3) | 1.0 (NA) | 1.0 (NA) |
| Master's | 62862 | 11.9 | 8.3 (8.1, 8.6) | 0.75 (0.72, 0.78) | 0.90 (0.87, 0.92) |
| Professional (e.g., MD, JD) | 14970 | 2.8 | 12.3 (11.6, 13.0) | 1.12 (1.05, 1.18) | 1.09 (1.04, 1.15) |
| Doctorate | 10969 | 2.1 | 23.9 (22.7, 25.1) | 2.16 (2.05, 2.28) | 1.20 (1.14, 1.25) |
| Missing | 70260 | 13.3 | 23.9 (23.5, 24.3) | 2.16 (2.10, 2.22) | 1.18 (1.10, 1.25) |
| Employment status | | | | | |
| Work outside home | 176197 | 33.3 | 21.2 (20.9, 21.4) | 2.48 (2.39, 2.57) | 1.33 (1.28, 1.37) |
| Work at home | 57246 | 10.8 | 8.5 (8.2, 8.8) | 1.0 (NA) | 1.0 (NA) |
| Does not work for pay | 223071 | 42.1 | 12.7 (12.5, 12.9) | 1.49 (1.43, 1.54) | 1.34 (1.29, 1.38) |
| Missing | 73144 | 13.8 | 23.9 (23.5, 24.3) | 2.80 (2.69, 2.91) | 1.33 (1.25, 1.41) |

Continued on next page

| | | | | | |
|---|---|---|---|---|---|
| US Region | | | | | |
| Midwest | 126686 | 23.9 | 18.1 (17.9, 18.4) | 1.50 (1.46, 1.55) | 1.10 (1.07, 1.13) |
| South | 182852 | 34.5 | 19.2 (19.0, 19.5) | 1.59 (1.55, 1.64) | 1.13 (1.10, 1.16) |
| Pacific | 73521 | 13.9 | 12.1 (11.7, 12.4) | 1.0 (NA) | 1.0 (NA) |
| Mountain | 42261 | 8.0 | 17.9 (17.4, 18.5) | 1.49 (1.43, 1.55) | 1.11 (1.07, 1.15) |
| Northeast | 88229 | 16.7 | 12.6 (12.3, 12.9) | 1.04 (1.01, 1.08) | 0.96 (0.93, 0.99) |
| Territories | 191 | <0.05 | 12.0 (6.3, 17.8) | 1.00 (0.52, 1.48) | 0.64 (0.44, 0.84) |
| Missing | 15918 | 3.0 | 33.3 (32.3, 34.3) | 2.76 (2.64, 2.87) | [c] |
| County urban classification | | | | | |
| Large central metro | 120722 | 22.8 | 11.7 (11.5, 12.0) | 1.0 (NA) | 1.0 (NA) |
| Large fringe metro | 115854 | 21.9 | 14.3 (14.0, 14.5) | 1.22 (1.18, 1.25) | 1.03 (1.01, 1.06) |
| Medium metro | 138457 | 26.1 | 16.8 (16.5, 17.1) | 1.43 (1.39, 1.47) | 1.13 (1.10, 1.16) |
| Small metro | 57778 | 10.9 | 21.0 (20.6, 21.5) | 1.79 (1.74, 1.85) | 1.18 (1.15, 1.22) |
| Micropolitan | 49266 | 9.3 | 24.2 (23.7, 24.7) | 2.06 (2.00, 2.12) | 1.19 (1.15, 1.23) |
| Non-core | 31472 | 5.9 | 27.4 (26.8, 28.1) | 2.34 (2.27, 2.41) | 1.23 (1.19, 1.27) |
| Missing | 16109 | 3.0 | 33.0 (32.0, 34.0) | 2.82 (2.71, 2.92) | [c] |

Juris Doctorate= JD; MD=Doctor of Medicine; NA=not applicable; NH=Non-Hispanic

[a] Race/ethnicity groups other than the group labeled “Hispanic” are non-Hispanic.

[b] Due to an interaction between age group and race/ethnicity, adjusted relative risks from the multivariable model are not reported in this table.

[c] Reliable estimates could not be calculated for the missing category for variables based on participants’ zip code, due to collinearity.

**sTable 9.** Sensitivity analysis: COVID-19 vaccine hesitancy in May 2021 political/by COVID-19 environment, health status, beliefs and behaviors among US adults, including those that self-described gender (N=529,658)

| | Sample | | COVID-19 vaccine hesitant | | |
|---|---|---|---|---|---|
| | **N** | **%** | **% (95% CI)** | **RR (95% CI)** | **Adj. RR (95% CI)** |
| State governor's political party | | | | | |
| Democratic | 282446 | 53.3 | 14.4 (14.2, 14.6) | 1.0 (NA) | [a] |
| Republican | 230264 | 43.5 | 19.2 (19.0, 19.4) | 1.33 (1.31, 1.36) | |
| Missing | 16948 | 3.2 | 31.8 (30.8, 32.7) | 2.21 (2.13, 2.28) | |
| County Trump vote total minus Biden vote total in 2020 presidential election | | | | | |
| Lowest quartile | 343255 | 64.8 | 12.8 (12.6, 12.9) | 1.0 (NA) | 1.0 (NA) |
| Second lowest quartile | 101627 | 19.2 | 21.9 (21.6, 22.3) | 1.72 (1.69, 1.75) | 1.27 (1.25, 1.30) |
| Second highest quartile | 47422 | 9.0 | 27.6 (27.0, 28.1) | 2.16 (2.11, 2.21) | 1.34 (1.30, 1.37) |
| Highest quartile | 19712 | 3.7 | 32.5 (31.7, 33.4) | 2.55 (2.48, 2.62) | 1.42 (1.38, 1.47) |
| Missing | 17642 | 3.3 | 32.4 (31.5, 33.4) | 2.54 (2.46, 2.62) | [b] |
| County COVID-19 April 2021 county death rate | | | | | |
| Lowest quartile | 26160 | 4.9 | 24.3 (23.7, 25.0) | 1.0 (NA) | 1.0 (NA) |
| Second lowest quartile | 168948 | 31.9 | 16.2 (15.9, 16.4) | 0.66 (0.64, 0.69) | 0.97 (0.94, 1.00) |
| Second highest quartile | 214630 | 40.5 | 15.7 (15.5, 15.9) | 0.65 (0.63, 0.67) | 1.00 (0.97, 1.03) |
| Highest quartile | 103804 | 19.6 | 17.1 (16.8, 17.4) | 0.70 (0.68, 0.73) | 1.01 (0.98, 1.04) |
| Missing | 16116 | 3.0 | 33.0 (32.0, 34.0) | 1.36 (1.30, 1.41) | [b] |
| Ever tested positive for COVID-19 | | | | | |
| Yes | 55851 | 10.5 | 20.7 (20.2, 21.1) | 1.24 (1.22, 1.27) | 1.10 (1.08, 1.13) |
| No or unsure | 470576 | 88.8 | 16.6 (16.5, 16.8) | 1.0 (NA) | 1.0 (NA) |
| Missing | 3231 | 0.6 | 19.6 (17.8, 21.4) | 1.18 (1.07, 1.29) | 0.94 (0.86, 1.01) |
| Ever diagnosed with high-risk medical condition | | | | | |
| One or more conditions | 324323 | 61.2 | 13.8 (13.6, 13.9) | 1.0 (NA) | 1.0 (NA) |
| No condition | 184503 | 34.8 | 19.4 (19.2, 19.7) | 1.41 (1.39, 1.43) | 1.01 (0.99, 1.02) |
| Missing | 20832 | 3.9 | 35.9 (35.0, 36.8) | 2.60 (2.53, 2.67) | 1.70 (1.65, 1.75) |
| Extent worried that you or someone in immediate family might become seriously ill from COVID-19 | | | | | |
| Worried | 209897 | 39.6 | 8.8 (8.6, 9.0) | 1.0 (NA) | 1.0 (NA) |
| Not too worried | 164794 | 31.1 | 13.7 (13.5, 13.9) | 1.55 (1.52, 1.59) | 1.31 (1.28, 1.35) |
| Not worried at all | 98919 | 18.7 | 33.7 (33.3, 34.1) | 3.82 (3.74, 3.91) | 1.78 (1.74, 1.83) |
| Missing | 56048 | 10.6 | 24.8 (24.3, 25.3) | 2.81 (2.74, 2.89) | 1.30 (1.11, 1.49) |
| Lives with someone or is 65 years or older | | | | | |
| Yes | 209290 | 39.5 | 12.6 (12.4, 12.8) | 1.0 (NA) | 1.0 (NA) |
| No | 204999 | 38.7 | 18.1 (17.9, 18.3) | 1.44 (1.41, 1.46) | 1.07 (1.04, 1.09) |
| No response | 115369 | 21.8 | 21.2 (20.9, 21.6) | 1.68 (1.65, 1.72) | 1.11 (1.08, 1.13) |
| Past-year flu vaccine | | | | | |
| Yes | 280787 | 53.0 | 5.6 (5.5, 5.7) | 1.0 (NA) | 1.0 (NA) |
| No or unsure | 193242 | 36.5 | 28.3 (28.0, 28.5) | 5.06 (4.94, 5.18) | 3.24 (3.16, 3.32) |
| Missing | 55629 | 10.5 | 24.9 (24.4, 25.4) | 4.46 (4.33, 4.59) | 2.12 (1.79, 2.45) |

Continued on next page

| | | | | | |
|---|---|---|---|---|---|
| Extent intentionally avoiding contact with others | | | | | |
| All of the time | 67156 | 12.7 | 11.0 (10.7, 11.3) | 1.0 (NA) | 1.0 (NA) |
| Most of the time | 142287 | 26.9 | 8.4 (8.2, 8.6) | 0.76 (0.73, 0.79) | 0.87 (0.84, 0.90) |
| Some of the time | 187201 | 35.3 | 9.0 (8.8, 9.1) | 0.81 (0.78, 0.84) | 0.88 (0.86, 0.91) |
| None of the time | 85930 | 16.2 | 44.5 (44.1, 45.0) | 4.03 (3.92, 4.15) | 2.43 (2.35, 2.50) |
| Missing | 47084 | 8.9 | 26.2 (25.6, 26.7) | 2.37 (2.29, 2.45) | 1.46 (1.37, 1.54) |

NA=not applicable

[a] State governor's political party was excluded from the multivariable model due to collinearity with county Trump vote share.

[b] Reliable estimates could not be calculated for the missing category for variables based on participants' zip code, due to collinearity.

**sTable 10**. Reasons for COVID-19 vaccine hesitancy in May 21 by race/ethnicity[a] among hesitant US adults (N=73362)[b]

| | White | Hispanic | Black | Asian | NA | PI | Multi-racial | Unknown |
|---|---|---|---|---|---|---|---|---|
| N | 44067 | 6028 | 3126 | 339 | 824 | 124 | 3117 | 15737 |
| | % (95% CI) | | | | | | | |
| Concerned about possible side effects | 51.5 (50.9, 52.1) | 49.3 (47.6, 50.9) | 51.4 (49.3, 53.5) | 48.5 (41.6, 55.5) | 41.4 (37.1, 45.7) | 43.7 (33.5, 53.9) | 58.5 (56.3, 60.7) | 42.0 (41.0, 43.0) |
| Don't trust COVID-19 vaccines | 50.5 (49.9, 51.0) | 44.8 (43.1, 46.5) | 46.1 (43.9, 48.2) | 38.4 (31.5, 45.4) | 45.7 (41.2, 50.1) | 45.0 (34.8, 55.1) | 55.1 (52.9, 57.3) | 44.8 (43.7, 45.8) |
| Don't trust the government | 43.2 (42.6, 43.8) | 35.9 (34.3, 37.6) | 34.3 (32.2, 36.3) | 26.2 (20.1, 32.2) | 42.9 (38.3, 47.5) | 34.4 (24.8, 44.1) | 50.0 (47.8, 52.3) | 40.0 (39.0, 41.0) |
| Don't believe I need it | 41.8 (41.2, 42.4) | 32.3 (30.6, 33.9) | 22.3 (20.4, 24.3) | 33.1 (26.2, 40.1) | 32.6 (28.6, 36.6) | 28.5 (18.9, 38.1) | 47.0 (44.7, 49.2) | 33.8 (32.9, 34.8) |
| Plan to wait and to see if safe | 33.9 (33.3, 34.5) | 41.8 (40.1, 43.4) | 41.1 (39.0, 43.2) | 42.5 (35.5, 49.5) | 27.0 (23.1, 30.8) | 35.5 (26.0, 45.1) | 34.4 (32.2, 36.5) | 31.2 (30.3, 32.2) |
| Concerned about an allergic reaction | 23.2 (22.7, 23.7) | 26.6 (25.1, 28.1) | 33.1 (31.0, 35.1) | 26.8 (20.0, 33.6) | 24.6 (21.0, 28.2) | 27.4 (18.5, 36.2) | 32.5 (30.4, 34.5) | 21.1 (20.3, 21.9) |
| Don't know if it will work | 22.1 (21.6, 22.6) | 22.7 (21.2, 24.1) | 21.2 (19.4, 23.0) | 26.4 (19.4, 33.4) | 17.2 (14.0, 20.3) | 24.7 (16.1, 33.4) | 27.6 (25.6, 29.6) | 20.1 (19.2, 20.9) |
| Don't like vaccines | 13.7 (13.3, 14.2) | 15.4 (14.0, 16.8) | 14.4 (12.9, 15.9) | 13.7 (9.5, 17.8) | 13.1 (10.5, 15.8) | 14.2 (7.2, 21.3) | 18.3 (16.5, 20.1) | 16.1 (15.3, 16.9) |
| Safety concern because of my health condition | 12.0 (11.7, 12.3) | 11.9 (10.7, 13.0) | 15.9 (14.5, 17.4) | 11.8 (8.0, 15.5) | 13.1 (10.8, 15.5) | 10.3 (4.4, 16.2) | 18.9 (17.3, 20.6) | 10.6 (10.1, 11.1) |
| Other people need it more | 11.9 (11.5, 12.3) | 16.9 (15.4, 18.4) | 10.4 (9.0, 11.7) | 17.6 (11.6, 23.6) | 10.6 (7.9, 13.2) | 15.5 (8.1, 22.8) | 15.7 (14.0, 17.5) | 12.3 (11.6, 13.0) |
| Doctor has not recommended | 9.0 (8.7, 9.3) | 9.7 (8.5, 11.0) | 7.6 (6.3, 8.9) | 7.7 (4.5, 10.8) | 9.5 (7.2, 11.8) | 2.8 (0.2, 5.4) | 14.7 (13.0, 16.3) | 9.2 (8.7, 9.8) |
| Against religious beliefs | 6.7 (6.4, 7.1) | 10.3 (9.1, 11.5) | 7.5 (6.4, 8.7) | 5.6 (3.1, 8.1) | 13.0 (10.3, 15.8) | 3.1 (0.4, 5.8) | 13.5 (12.0, 15.0) | 9.7 (9.1, 10.3) |
| Currently/planning to be pregnant/breastfeeding | 6.1 (5.8, 6.4) | 10.0 (8.8, 11.2) | 7.0 (5.9, 8.1) | 11.4 (7.5, 15.3) | 4.1 (2.3, 5.8) | 4.1 (0.4, 7.9) | 8.5 (7.2, 9.8) | 6.2 (5.7, 6.6) |
| Concerned about cost | 2.3 (2.1, 2.5) | 4.9 (3.8, 6.1) | 3.3 (2.5, 4.0) | 3.9 (-0.2, 8.0) | 2.1 (1.0, 3.2) | 2.5 (-0.5, 5.4) | 5.5 (4.4, 6.6) | 4.3 (3.9, 4.8) |
| Other | 15.2 (14.8, 15.6) | 13.2 (12.2, 14.2) | 11.5 (10.2, 12.9) | 16.8 (10.6, 23.0) | 19.5 (16.2, 22.7) | 17.9 (10.3, 25.6) | 22.9 (21.0, 24.8) | 19.0 (18.3, 19.8) |

NA=Native American; NH=Non-Hispanic; PI=Pacific Islander.

[a] Race/ethnicity groups other than the group labeled “Hispanic” are non-Hispanic.

[b] Answered that they probably or definitely would not choose to get vaccinated if offered a vaccine to prevent COVID-19.
